# Vitamin D Deficiency, Supplementation, and Aging-Related Morbidity and Mortality Across Two Longitudinal Population Cohorts

**DOI:** 10.1101/2025.05.29.25328548

**Authors:** Ariel Israel, Abraham Weizman, Sarah Israel, Shai Ashkenazi, Eytan Ruppin, Eli Magen, Eugene Merzon, Shlomo Vinker

**Affiliations:** Leumit Research Institute, Leumit Health Services; Tel-Aviv, Israel; Department of Epidemiology and Preventive Medicine, School of Public Health, Faculty of Medical & Health Sciences, Tel Aviv University; Tel-Aviv, Israel; Research Unit, Geha Mental Health Center, Felsenstein Medical Research Center and Sagol School of Neurosciences, Faculty of Medical & Health Sciences, Tel-Aviv University, Tel-Aviv, Israel; Department of Clinical Microbiology and Infectious Diseases and Department of Internal Medicine, Hadassah-Hebrew University Medical Center, Faculty of Medicine, Hebrew University of Jerusalem, Jerusalem Israel; Adelson School of Medicine, Ariel University; Ariel, Israel; Schneider Children’s Medical Center, Petah-Tikva, Israel; Translational Research Institute and the Department of Surgery, Cedars-Sinai Medical Center, Los Angeles, California, USA; Medicine A Department, Assuta Ashdod University Medical Center, Ben Gurion University of the Negev; Beer Sheva, Israel

**Keywords:** Vitamin D Deficiency, Aging, Mortality, Healthy Aging, Electronic Health Records, Cohort Studies, Causal Inference

## Abstract

Vitamin D deficiency has been implicated in biological processes linked to aging and organismal resilience, yet its relationship to long-term systemic aging trajectories remains incompletely characterized. We analyzed longitudinal electronic health record data from two large healthcare systems: Leumit Health Services (LHS), a nationwide Israeli health organization, and TriNetX, a federated US research network containing de-identified electronic health records from over 120 million individuals.

We examined more than 1.67 million serum 25-hydroxyvitamin D [25(OH)D] measurements from over 468,500 adults in LHS (2009-2020). Longitudinal matched analyses were performed in LHS, and associations involving severe deficiency were externally validated in 223,000 propensity score-matched pairs from TriNetX.

In both populations, severe deficiency was associated with increased risks across metabolic, cardiovascular, neurodegenerative, renal, microvascular, and mortality-related outcomes, including diabetes mellitus, myocardial infarction, cerebrovascular disease, dementia, dialysis, diabetic retinopathy, and foot/toe amputation. To assess the potential health benefits and modifiability of supplementation, we modeled vitamin D supplementation longitudinally using pharmacy-dispensed purchases calibrated to predict serum 25(OH)D changes over time. Supplementation was independently associated with dose-dependent reductions across mortality and multiple aging-related outcomes, while showing no corresponding protective association for skin malignancy, a negative-control outcome strongly linked to ultraviolet exposure.

To strengthen causal interpretation, we additionally implemented inverse-probability-of-treatment-weighted marginal structural Cox models for mortality, which yielded results consistent with, and in several cases stronger than, conventional adjusted models.

Together, these findings support severe vitamin D deficiency as an important and potentially modifiable determinant of systemic aging vulnerability and age-related multimorbidity in human populations.

## Introduction

Vitamin D deficiency is widespread worldwide, particularly in populations with limited sun exposure or darker skin pigmentation (Hilger et al. 2014; Holick 2007; Cashman et al. 2016). While its role in bone health is well established, increasing evidence has linked low serum 25-hydroxyvitamin D [25(OH)D] levels to increased mortality (Gotsman et al. 2012) and a wide range of chronic conditions, including diabetes (Scragg et al. 1995; Pittas et al. 2007), cardiovascular disease(Judd & Tangpricha 2009), retinopathy, chronic kidney disease, foot ulcers, and neurodegenerative disorders (Usluogullari et al. 2015; Liu et al. 2011; Liu et al. 2024; Afzal et al. 2013; Vasdeki et al. 2024). However, despite extensive observational research, the causality of these associations remains disputed. One major challenge is disentangling whether low vitamin D levels are a contributing factor to poor health, or merely a biomarker of existing disease or adverse health behaviors.

Israel, like many countries, has experienced persistently high rates of diabetes-related complications, including lower-extremity amputations, with rates particularly elevated compared to other OECD nations(Anon n.d.; OECD Reviews of Health Care Quality 2012). At the same time, vitamin D deficiency is common in several demographic subgroups, particularly among individuals with limited sun exposure due to lifestyle, occupational, or cultural practices. Yet, large-scale longitudinal studies explicitly quantifying the long-term impact of severe vitamin D deficiency on aging-related outcomes, and evaluating whether such risks are modifiable, remain scarce.

In this study, we leveraged the electronic health records (EHRs) of two large, demographically distinct populations—Leumit Health Services (LHS) in Israel and the US-based TriNetX Research Network—to assess the long-term clinical impact of severe vitamin D deficiency (<10 ng/mL) versus sufficiency (>30 ng/mL). Using tightly matched cohorts, we examined risks of mortality, diabetes, microvascular and macrovascular complications, and dementia, representing multiple major domains of age-related morbidity. We further replicated findings across both healthcare systems for robustness and evaluated whether pharmacy-documented vitamin D supplementation conferred both independent and dose-dependent risk reduction after adjusting for baseline 25(OH)D levels. We further applied causal-inference analyses using inverse-probability-of-treatment-weighted marginal structural Cox models to evaluate whether these associations remained robust after accounting for time-varying treatment selection and measured confounding.

Our study is based on four sequential analyses (Figure 1):

**Analysis A:** We first explored the distribution of 25(OH)D in the LHS population using >1.67 million measurements from >470,000 individuals, modeling seasonal variation and the effect of vitamin D products on serum levels.
**Analysis B:** We evaluated the impact of severe vitamin D deficiency by following a matched LHS cohort of 12,352 severely deficient versus sufficient individuals, matched on age, sex, BMI, smoking status, blood pressure, socioeconomic status, and geodemographic classification, using longitudinal time-to-event models to assess mortality and vitamin D-deficiency complications.
**Analysis C:** We validated these associations in the US-based TriNetX network by conducting an analogous analysis in matched cohorts of 206,600 individuals with severe deficiency and 206,600 with sufficient levels for the same outcomes.
**Analysis D:** Finally, we assessed the potential of vitamin D supplementation to mitigate these risks in LHS by fitting time-dependent Cox proportional hazards models incorporating pharmacy-dispensed vitamin D supplementation as a time-varying covariate together with baseline 25(OH)D levels. To strengthen causal interpretation and reduce bias from time-varying treatment selection, we additionally implemented inverse-probability-of-treatment-weighted marginal structural Cox models for mortality.

**Figure 1:**
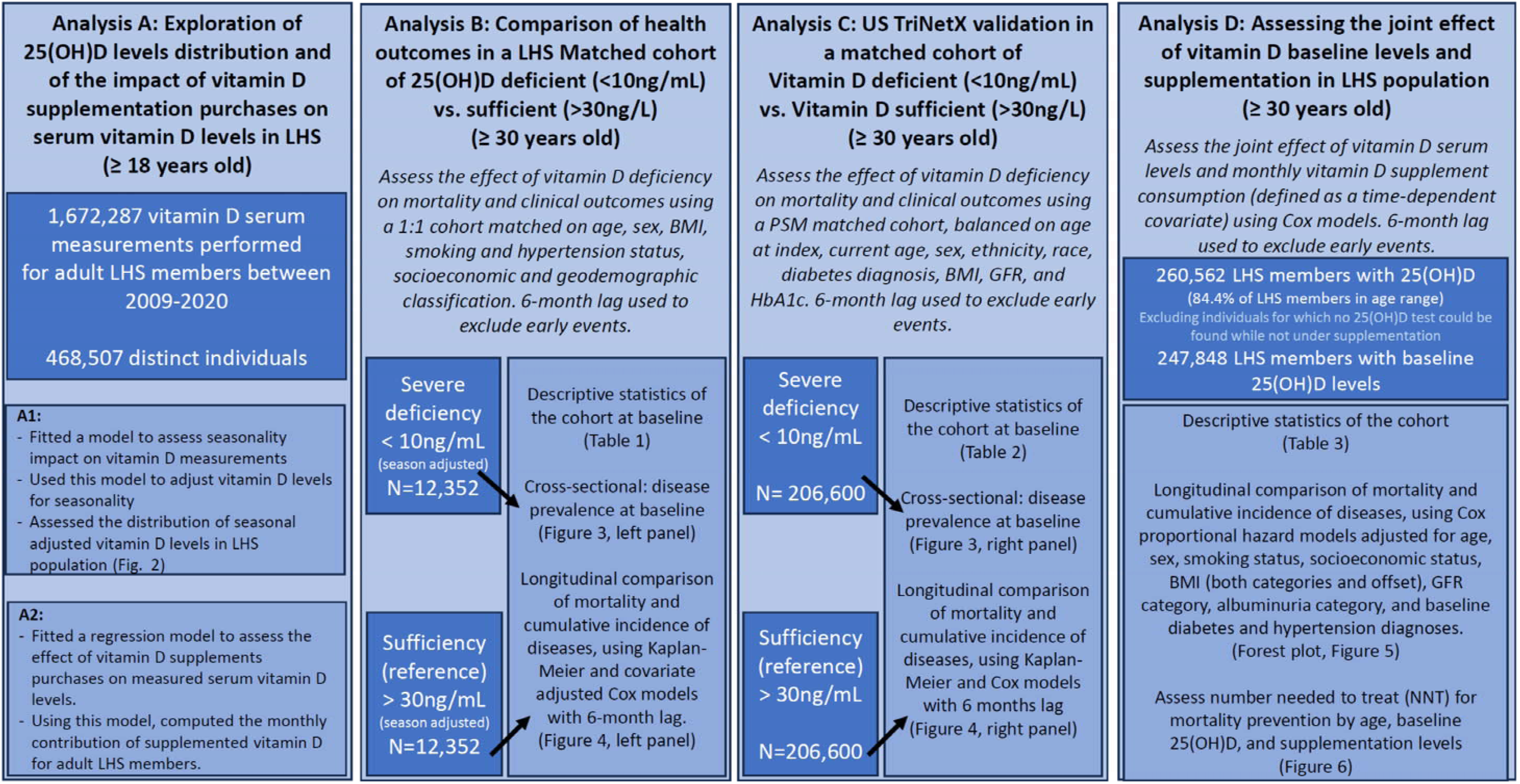
Overview of the analyses performed, showing study populations, age thresholds, exposure definitions, matching, and statistical models used.

**Figure 2:**
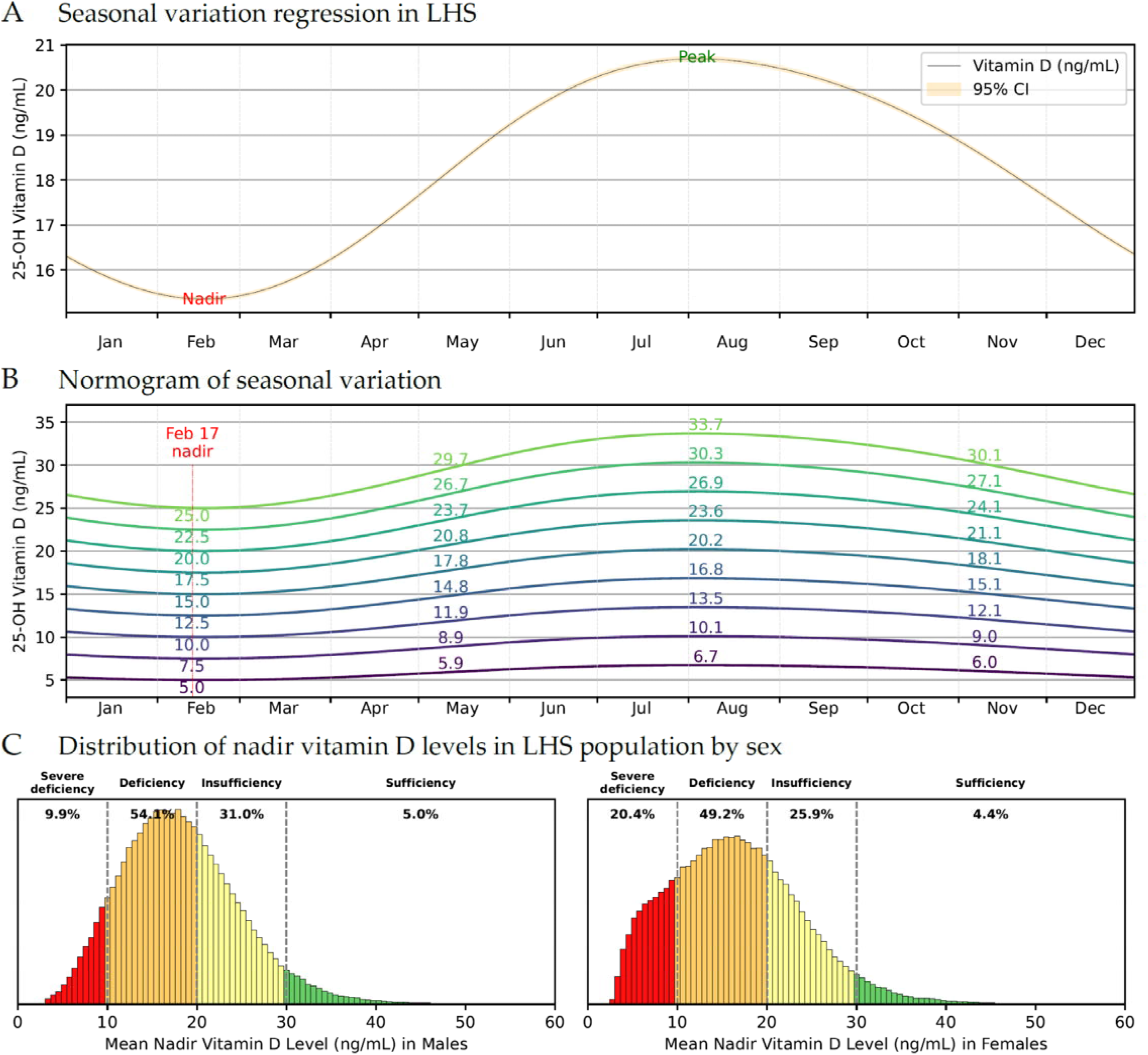
Seasonal variation and population distribution of serum 25(OH)D in LHS (Analysis A) **(A)** Modeled seasonal fluctuation in serum 25(OH)D levels across the calendar year in the Leumit Health Services (LHS) population, showing a sinusoidal pattern with a nadir in mid-February and peak in late August. The 95% confidence interval is shown but appears nearly indistinguishable due to the large population size. **(B)** Normogram translating observed 25(OH)D levels to their expected nadir values based on date of measurement, enabling standardization of vitamin D status across seasons. **(C)** Sex-stratified distribution of mean nadir-adjusted vitamin D levels in the LHS population. Severe deficiency (<10 ng/mL) is substantially more common in females (20.4%) than in males (9.9%), whereas vitamin D sufficiency (>30 ng/mL) is uncommon in both sexes.

## Materials and Methods

### Overview

We conducted a retrospective observational study across two healthcare systems (Israel: Leumit Health Services, LHS; United States: TriNetX). Analyses were prespecified and categorized as cross-sectional (baseline prevalence) or longitudinal (time-to-event). Figure 1 summarizes analytic populations, eligibility, exposure definitions, matching/PSM, and lags.

### Participants

- Analysis A (LHS): adults ≥18 years with ≥1 serum 25(OH)D test in 2009–2020 (index date = first qualifying test).
- Analyses B–D (LHS): adults ≥30 years (index = first qualifying test); a 6-month lag excluded events within the first year after index. Sensitivity analyses were also repeated with a 12-month lag, yielding similar results (Supplementary Figure 2).
- Analysis C (TriNetX): adults ≥18 years, with harmonized outcome definitions; propensity score matching (PSM) applied as below.

Full inclusion/exclusion criteria are detailed in Figure 1.

### Variables

#### Covariates (LHS)

Cox models were fitted after adjustment for age, sex, smoking status, socioeconomic status, BMI (fine categories plus within-category offset), eGFR category, albuminuria category, baseline diabetes (ICD-9 diagnosis or HbA1c >=6.5%), baseline chronic kidney disease, hypertension, and systolic blood pressure. A 6-month lag excluded early events to reduce reverse causation.

#### Covariates (TriNetX)

we used propensity score matching (PSM) using age, sex, ethnicity, race, diabetes diagnosis, BMI, eGFR, and hemoglobin A1c categories; a 6-month lag excluded early events.

Outcomes included all-cause mortality and clinical outcomes (myocardial infarction, heart failure, dialysis, diabetic retinopathy, foot/toe amputation, dementia, skin malignancy, and others), as defined in Supplementary Table 4.

### Data Sources and Measurement

We used de-identified electronic health records (EHRs) from two distinct cohorts: Leumit Health Services (LHS), a nationwide Israeli health maintenance organization, and the TriNetX US Collaborative Network, which aggregates de-identified EHRs from over 70 healthcare organizations (HCOs). LHS and TriNetX EHR sources included demographics, diagnoses (ICD-9/ICD-10 as applicable), laboratory measurements, anthropometric measures, pharmacy dispensing, and mortality data.

Serum 25(OH)D levels were reported in ng/mL units (1 ng/mL = 2.5 nmol/L), and were calendar-adjusted for seasonality in LHS (see Seasonal Normalization). Medical conditions were defined using diagnosis-code groups and laboratory thresholds. An exploratory outcome-discovery analysis (Analysis B) screened approximately 300 curated medical-condition groups developed at the Leumit Research Institute and subsequently validated significant associations in TriNetX. The TriNetX platform enables retrospective longitudinal analyses using standardized temporal alignment and harmonized outcome definitions.

### Bias and sensitivity analyses

We mitigated confounding using several complementary approaches: (i) exact matching and multivariable-adjusted Cox models in LHS analyses, (ii) propensity score matching with balance diagnostics in TriNetX, and (iii) time-dependent supplementation analyses incorporating longitudinal pharmacy dispensing data. To reduce reverse causation, prevalent events were excluded and longitudinal analyses applied a 6-month lag, with additional sensitivity analyses using a 12-month lag.

To strengthen causal interpretation of the supplementation analyses, we additionally performed a causal-framework sensitivity analysis using inverse-probability of treatment weighted (IPTW) marginal structural Cox models (MSM) for all-cause mortality, the primary endpoint. Stabilized treatment weights were estimated using multinomial logistic regression models predicting observed time-updated supplementation categories conditional on prior supplementation status and measured baseline covariates, including baseline serum 25(OH)D category, age, sex, smoking status, socioeconomic status, BMI category, BMI within-category offset, baseline diabetes, chronic kidney disease, hypertension, systolic blood pressure, eGFR category, and albuminuria category.

For each individual-month interval, stabilized weights were calculated as the ratio between the probability of the observed supplementation category conditional on prior supplementation history alone (numerator model) and the probability conditional on both supplementation history and measured confounders (denominator model). Weighted Cox proportional hazards models with robust variance estimation clustered by individual were then fitted using these stabilized interval-level weights. These analyses were intended to reduce bias from time-varying treatment selection and measured confounding in longitudinal supplementation analyses.

### Seasonal Normalization of Serum 25(OH)D levels

25(OH)D levels display marked seasonal variation. To standardize values across measurement dates, we fitted a sinusoidal regression model using log-transformed 25(OH)D values with first-and second-order sine and cosine terms to model annual periodicity:

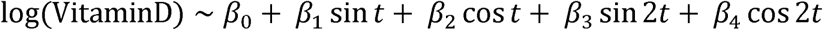

where t denotes the day of the year in radians. Adjusted values were computed by estimating serum levels at the population trough (February 14), using the fitted model to remove seasonal effects.

### Data Extraction and Preprocessing

Structured EHR data from LHS were queried using T-SQL. Python 3.11 and the Pandas library were used for data preprocessing. Each patient record was anonymized using an internal study identifier. Baseline covariates, including sex, age, BMI category, socioeconomic status, ethnic sector, smoking status, kidney function, albuminuria, and blood pressure were extracted as of the index date.

### Matched Cohort Construction (LHS)

To evaluate the long-term impact of severe deficiency, we constructed a 1:1 matched cohort comparing individuals with calendar-adjusted vitamin D levels <10 ng/mL and those with >30 ng/mL. Matching was exact on age category, sex, smoking status, BMI category, blood-pressure category, ethnic sector (general, Arab, ultra-Orthodox), socioeconomic status, and year of LHS enrollment, ensuring balance in major demographic and metabolic risk factors. Baseline tables and models report groups by vitamin D status categories; the sufficiency group (>30 ng/mL) served as the reference category.

### Outcome Discovery in LHS

Approximately 300 common medical conditions, defined using validated ICD-9 code groups curated by the Leumit Research Institute, were screened in the matched cohort. Cox proportional hazards models were used to compare cumulative incidence between groups, adjusting for covariates. False discovery rate (FDR) control was applied to identify conditions with significant differences.

### TriNetX Validation Cohort

For external validation, we compared TriNetX patients with 25(OH)D <10 ng/mL to those with >30 ng/mL. Propensity score matching was performed on age, sex, African American ethnicity, BMI category, obesity diagnosis, smoking status, diabetes diagnosis, eGFR category, and hemoglobin A1c category. Post-matching balance was assessed using standardized mean differences. Conditions identified as FDR-significant in LHS were validated using harmonized definitions in TriNetX with a 6-month lag to exclude early events.

### Vitamin D supplementation estimation

All vitamin D-containing product purchases were extracted from Leumit Health Services pharmacy records. Each product in the catalog was assigned a total vitamin D content (expressed in kilo-international units, kIU) based on labeled concentration and number of dosage units per package.

To quantify the contribution of supplementation to serum 25-hydroxyvitamin D [25(OH)D] levels, we implemented a two-stage regression framework:

1. **Baseline Model**: A multivariable linear regression was used to predict log-transformed serum 25(OH)D levels based on demographic and structural variables, including age, sex, BMI category, ethnicity, socioeconomic status, and seasonal harmonics (sine and cosine terms for date). This model explained 25.4% of the variance in serum 25(OH)D levels.
2. **Supplementation Model**: The residuals from the baseline model were regressed on lagged vitamin D supplement purchases. Total purchased vitamin D (in kIU) was aggregated across time bins—1 month, 3 months (quarter), and 12 months (year) preceding the test date. Each time bin was assigned an empirically estimated coefficient reflecting its weighted contribution to the serum level at the target date.

### Validation of Supplementation Model and Avoidance of Reverse Causation

Although clinical factors such as impaired renal function may influence vitamin D metabolism, no clinical or demographic variables were included in the supplementation model itself to minimize potential reverse causation. The baseline model explained 25.4% of total variance in serum 25(OH)D levels, while the supplementation model explained an additional 9.3% of residual variance. Together, the combined framework explained approximately 35% of total variance in serum 25(OH)D, supporting the measurable dose-dependent contribution of documented supplementation.

### Classification of Vitamin D Status

Baseline vitamin D status was defined using each individual’s first calendar-adjusted serum 25(OH)D measurement and grouped into categories: 0-5 ng/mL, 5-10 ng/mL, 10-15 ng/mL, 15-20 ng/mL, and >20 ng/mL (reference category).

For each individual and each follow-up interval, the estimated serum contribution of vitamin D supplementation was calculated from pharmacy dispensing records and expressed in ng/mL-equivalent categories: 0-0.2 ng/mL (reference), 0.2-5 ng/mL, 5-10 ng/mL, and >10 ng/mL. This time-varying supplementation exposure was incorporated into Cox proportional hazards models.

### Survival Analysis Using Time-Dependent Supplementation

We used time-dependent Cox proportional hazards models to evaluate supplementation effects on mortality and clinical outcomes. Person-time was divided into longitudinal follow-up intervals per patient, updating supplementation exposure over time using pharmacy dispensing records. Models jointly included baseline 25(OH)D categories and time-dependent supplementation categories and were adjusted for demographic, metabolic, renal, and cardiovascular covariates.

In MSM/IPTW sensitivity analyses, weighted Cox proportional hazards models were fitted using cumulative stabilized inverse-probability weights to estimate associations under a pseudo-population in which measured confounding of supplementation assignment was reduced. Robust sandwich variance estimation clustered by individual was used in all weighted analyses.

### Absolute Risk Reduction and Number Needed to Treat (NNT)

Absolute risk reduction (ARR) and number needed to treat (NNT) were estimated at10 years:

1. **Stratification**: Individuals were grouped by baseline vitamin D status.
2. **Time-Dependent Survival Modeling**: Survival probabilities were estimated from adjusted Cox proportional hazards models using time-updated supplementation exposure.
3. **Standardized Reference Profile**: A representative reference profile was generated using average continuous covariates and modal categorical covariates from the reference supplementation group (<0.2 ng/mL equivalent supplementation).
4. **Survival Projection**: Standardized survival curves were estimated under each supplementation category.
5. ARR and NNT Calculation:

- ARR(t) = S1(t) - S0(t)
- NNT(t) = 1 / |ARR(t)|

Additional IPTW/MSM-based ARR and NNT estimates were calculated for mortality as a causal-framework sensitivity analysis using the weighted marginal structural Cox model.

#### Statistical Analyses

Categorical variables were compared using Fisher’s exact test. Continuous variables were compared using two-sample t-tests. Logistic regression was used to estimate odds ratios (ORs) with 95% confidence intervals (CIs). Longitudinal outcomes were analyzed using Cox proportional hazards models. Kaplan-Meier curves were used to visualize survival. Death and disenrollment were treated as censoring events in all models. Given the strong association between vitamin D deficiency and mortality itself, competing-risk models would likely underestimate rather than inflate relative hazards for nonfatal outcomes.

All analyses were performed using R 4.4 and Python 3.11.

### Generative AI statement

ChatGPT was used to assist with language refinement for improved clarity and flow, and to provide support with coding tasks related to data analysis scripts. All content was reviewed and verified by the authors to ensure accuracy and integrity.

### Ethics

All analyses of Leumit Health Services (LHS) data were performed retrospectively on fully anonymized electronic health records under institutional review board approval (Leumit Health Services IRB approval no. LEU-0020-24), with a waiver of informed consent granted because the study posed minimal risk and used de-identified data.

Analyses using the US TriNetX Research Network were conducted on de-identified aggregate data provided under data-use agreements with participating healthcare organizations and were therefore not considered human-subjects research; these analyses were compliant with the US Health Insurance Portability and Accountability Act (HIPAA) and local institutional guidelines.

### Data Availability

Individual-level patient data from LHS are not publicly available due to privacy and regulatory restrictions. Aggregated data supporting the findings of this study may be provided to qualified researchers upon reasonable request, subject to approval by the LHS Institutional Review Board.

Data from the TriNetX network are available to institutions that are authorized by the TriNetX collaboration network, through direct request to TriNetX (https://www.trinetx.com).

## Results

### Analysis A: Exploration of vitamin D distribution and of the impact of vitamin D supplementation purchases on serum 25(OH)D levels in LHS

We began our analysis with an exploratory assessment of serum 25(OH)D test results collected between 2009 and 2020 from LHS members aged 18 years or older. Serum 25(OH)D levels demonstrated pronounced seasonal variation, consistent with fluctuations in sun exposure throughout the year. To model this variation, we applied a sinusoidal regression incorporating first- and second-order sine and cosine terms to log-transformed 25(OH)D levels. The fitted model revealed a robust periodic signal (see Supplementary Table 1), with peak vitamin D levels observed in late summer and a nadir around mid-February (Figure 1A).

To normalize for seasonality, we derived a population-based normogram (Figure 1B) that maps measured vitamin D levels to their expected nadir equivalents. This calendar-adjustment procedure was applied to all serum measurements, enabling generation of a seasonally adjusted annual exposure metric for each measured vitamin D levels. We then computed the average of these calendar-adjusted levels per person to reflect long-term vitamin D status. The resulting distribution (Figure 1C) highlights a substantial prevalence of severe deficiency (<10 ng/mL) and a smaller subgroup with sufficient average levels (>30 ng/mL). Severe deficiency was substantially more common in females, a pattern that aligns with known differences in sun exposure behaviors among women in certain cultural and religious communities within the population.

### Analysis B: LHS Matched cohort of 25(OH)D deficient (<10ng/mL) vs. sufficient individuals - baseline

To evaluate the clinical consequences of severe vitamin D deficiency, we constructed a matched cohort comparing individuals with severe deficiency to those with sufficiency (flowchart in Supplementary Figure 1). The index date was defined as the first recorded serum 25(OH)D measurement for each individual from 2009 onward. Using the previously fitted seasonal-correction model, we calculated a day-of-year–adjusted mean 25(OH)D level for each individual based on the index measurement and classified participants into two exposure groups: severely deficient (<10 ng/mL) and sufficient (>30 ng/mL).

For each severely deficient individual, we selected a matched control using 1:1 exact matching on age category, sex, smoking status, body-mass-index (BMI) category, hypertension status, socioeconomic status (SES), and geodemographic classification (general population, Arab, or Ultra-Orthodox) at the index date; among eligible controls, the individual with the closest BMI was selected.

Table 1 presents the baseline characteristics of the matched cohort, comprising 12,352 individuals with severe vitamin D deficiency (mean 25(OH)D < 10 ng/mL) and 12,352 matched controls with sufficient levels (> 30 ng/mL at baseline).

**Table 1.**
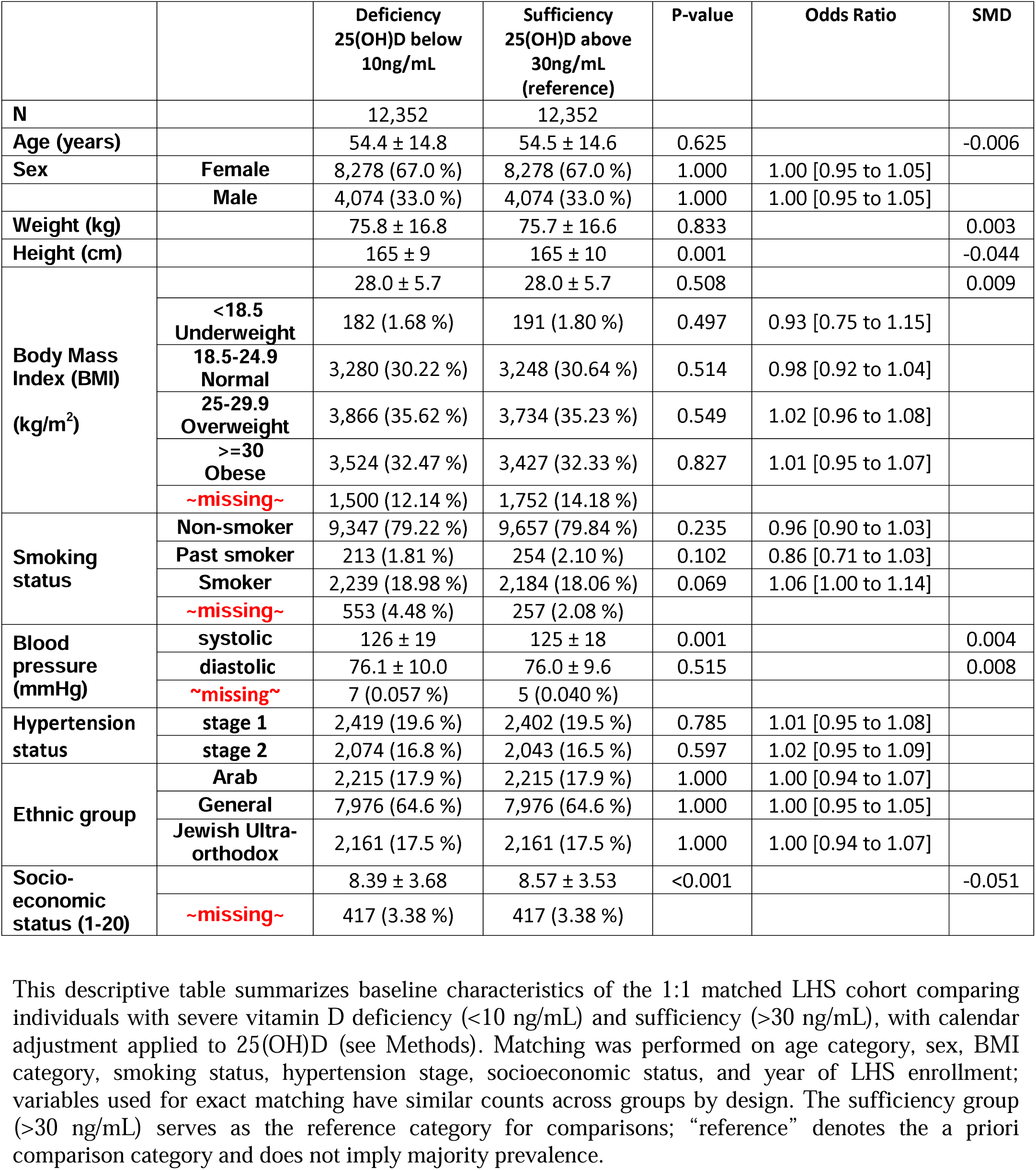
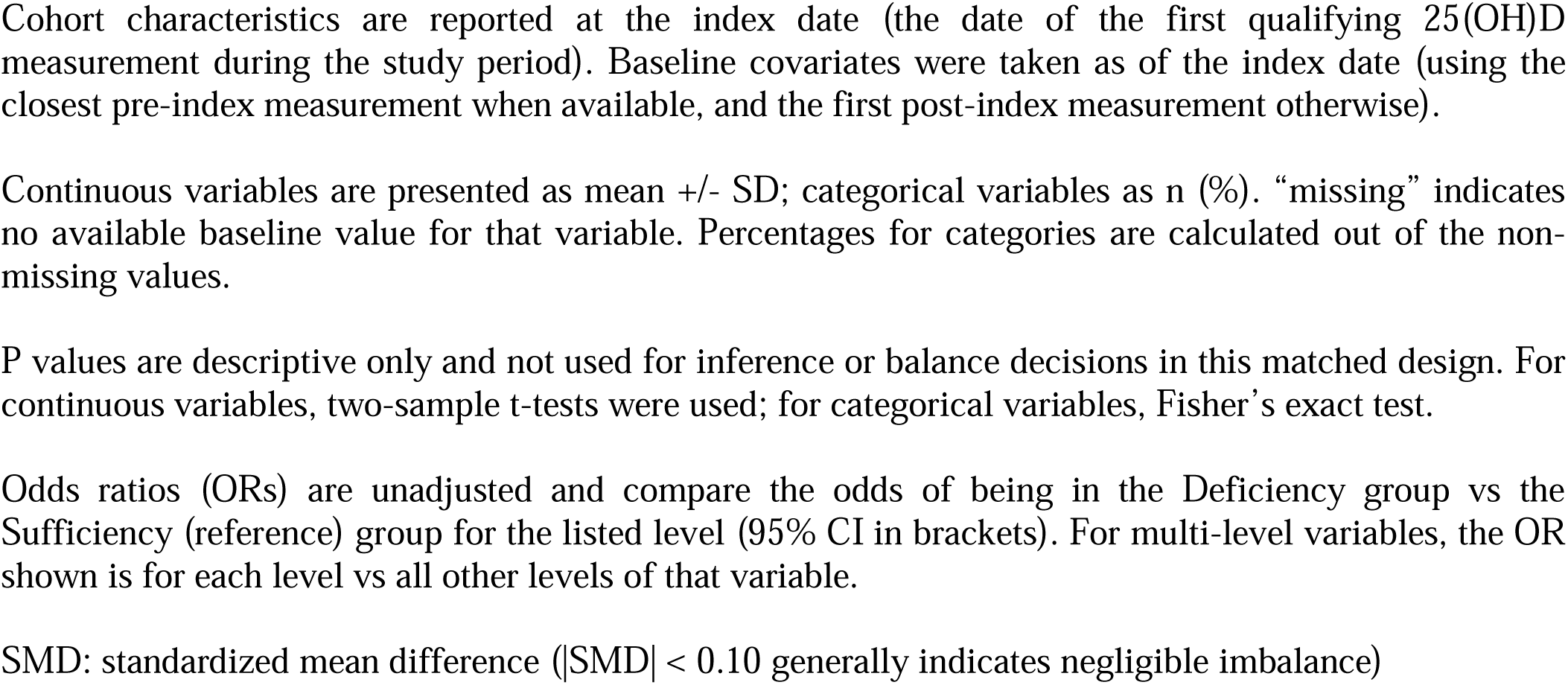
Demographic and clinical characteristics of the LHS matched cohort at baseline (Analysis B)

|  |  | Deficiency<br>25(OH)D below<br>10ng/mL | Sufficiency<br>25(OH)D above<br>30ng/mL<br>(reference) | P-value | Odds Ratio | SMD |
| --- | --- | --- | --- | --- | --- | --- |
| **N** |  | 12,352 | 12,352 |  |  |  |
| **Age (years)** |  | 54.4 ± 14.8 | 54.5 ± 14.6 | 0.625 |  | -0.006 |
| **Sex** | **Female** | 8,278 (67.0 %) | 8,278 (67.0 %) | 1.000 | 1.00 [0.95 to 1.05] |  |
|  | **Male** | 4,074 (33.0 %) | 4,074 (33.0 %) | 1.000 | 1.00 [0.95 to 1.05] |  |
| **Weight (kg)** |  | 75.8 ± 16.8 | 75.7 ± 16.6 | 0.833 |  | 0.003 |
| **Height (cm)** |  | 165 ± 9 | 165 ± 10 | 0.001 |  | -0.044 |
| **Body Mass Index (BMI) (kg/m<sup>2</sup>)** |  | 28.0 ± 5.7 | 28.0 ± 5.7 | 0.508 |  | 0.009 |
|  | **<18.5 Underweight** | 182 (1.68 %) | 191 (1.80 %) | 0.497 | 0.93 [0.75 to 1.15] |  |
|  | **18.5-24.9 Normal** | 3,280 (30.22 %) | 3,248 (30.64 %) | 0.514 | 0.98 [0.92 to 1.04] |  |
|  | **25-29.9 Overweight** | 3,866 (35.62 %) | 3,734 (35.23 %) | 0.549 | 1.02 [0.96 to 1.08] |  |
|  | **>=30 Obese** | 3,524 (32.47 %) | 3,427 (32.33 %) | 0.827 | 1.01 [0.95 to 1.07] |  |
|  | **~missing~** | 1,500 (12.14 %) | 1,752 (14.18 %) |  |  |  |
| **Smoking status** | **Non-smoker** | 9,347 (79.22 %) | 9,657 (79.84 %) | 0.235 | 0.96 [0.90 to 1.03] |  |
|  | **Past smoker** | 213 (1.81 %) | 254 (2.10 %) | 0.102 | 0.86 [0.71 to 1.03] |  |
|  | **Smoker** | 2,239 (18.98 %) | 2,184 (18.06 %) | 0.069 | 1.06 [1.00 to 1.14] |  |
|  | **~missing~** | 553 (4.48 %) | 257 (2.08 %) |  |  |  |
| **Blood pressure (mmHg)** | **systolic** | 126 ± 19 | 125 ± 18 | 0.001 |  | 0.004 |
|  | **diastolic** | 76.1 ± 10.0 | 76.0 ± 9.6 | 0.515 |  | 0.008 |
|  | **~missing~** | 7 (0.057 %) | 5 (0.040 %) |  |  |  |
| **Hypertension status** | **stage 1** | 2,419 (19.6 %) | 2,402 (19.5 %) | 0.785 | 1.01 [0.95 to 1.08] |  |
|  | **stage 2** | 2,074 (16.8 %) | 2,043 (16.5 %) | 0.597 | 1.02 [0.95 to 1.09] |  |
| **Ethnic group** | **Arab** | 2,215 (17.9 %) | 2,215 (17.9 %) | 1.000 | 1.00 [0.94 to 1.07] |  |
|  | **General** | 7,976 (64.6 %) | 7,976 (64.6 %) | 1.000 | 1.00 [0.95 to 1.05] |  |
|  | **Jewish Ultra-orthodox** | 2,161 (17.5 %) | 2,161 (17.5 %) | 1.000 | 1.00 [0.94 to 1.07] |  |
| **Socio-economic status (1-20)** |  | 8.39 ± 3.68 | 8.57 ± 3.53 | <0.001 |  | -0.051 |
|  | **~missing~** | 417 (3.38 %) | 417 (3.38 %) |  |  |  |

This descriptive table summarizes baseline characteristics of the 1:1 matched LHS cohort comparing individuals with severe vitamin D deficiency (<10 ng/mL) and sufficiency (>30 ng/mL), with calendar adjustment applied to 25(OH)D (see Methods). Matching was performed on age category, sex, BMI category, smoking status, hypertension stage, socioeconomic status, and year of LHS enrollment; variables used for exact matching have similar counts across groups by design. The sufficiency group (>30 ng/mL) serves as the reference category for comparisons; “reference” denotes the a priori comparison category and does not imply majority prevalence.
Cohort characteristics are reported at the index date (the date of the first qualifying 25(OH)D measurement during the study period). Baseline covariates were taken as of the index date (using the closest pre-index measurement when available, and the first post-index measurement otherwise).
5 Continuous variables are presented as mean +/- SD; categorical variables as n (%). “missing” indicates no available baseline value for that variable. Percentages for categories are calculated out of the non-missing values.
10 P values are descriptive only and not used for inference or balance decisions in this matched design. For continuous variables, two-sample t-tests were used; for categorical variables, Fisher’s exact test.
Odds ratios (ORs) are unadjusted and compare the odds of being in the Deficiency group vs the Sufficiency (reference) group for the listed level (95% CI in brackets). For multi-level variables, the OR shown is for each level vs all other levels of that variable.
15 SMD: standardized mean difference ( $|SMD| < 0.10$ generally indicates negligible imbalance)

As expected from the matching design, the two groups were well balanced across all matching variables, with negligible standardized mean differences (SMD < 0.01 for age, BMI, weight and blood pressure measures). The matched groups (mean age 54.4 years; 67% female) showed no meaningful differences in the matched variables. Overall, the baseline risk factors in the two groups at the time of serum 25(OH)D testing were highly similar, supporting comparability between groups and strengthening the validity of the subsequent risk-association analyses.

We examined baseline clinical associations with severe vitamin D deficiency in the LHS matched cohort (Figure 3A). Despite rigorous matching on demographic and metabolic factors, individuals with severe deficiency showed markedly higher baseline prevalence of chronic conditions. Elevated odds were observed for diabetes-related and cardiovascular outcomes, including foot/toe amputation (OR 4.01, 95% CI [2.04 to 8.62], p<10^-5^), diabetes mellitus (OR 1.23, 95% CI [1.15 to 1.31], p<10^-9^), diabetic retinopathy (OR 1.69, 95% CI [1.45 to 1.99], p<10^-10^), dementia (OR 1.89, 95% CI [1.53 to 2.33], p<10^-9^), cerebrovascular accident (CVA) (OR 1.79, 95% CI [1.50 to 2.14], p<10^-8^), and myocardial infarction (OR 1.40, 95% CI [1.25 to 1.57], p<10^-8^). In contrast, the odds of skin malignancy were markedly reduced (OR 0.44, 95% CI [0.31 to 0.61], p<10^-6^), suggesting lower cumulative skin ultraviolet (UV) exposure in the group with low 25(OH)D serum levels.

**Figure 3:**
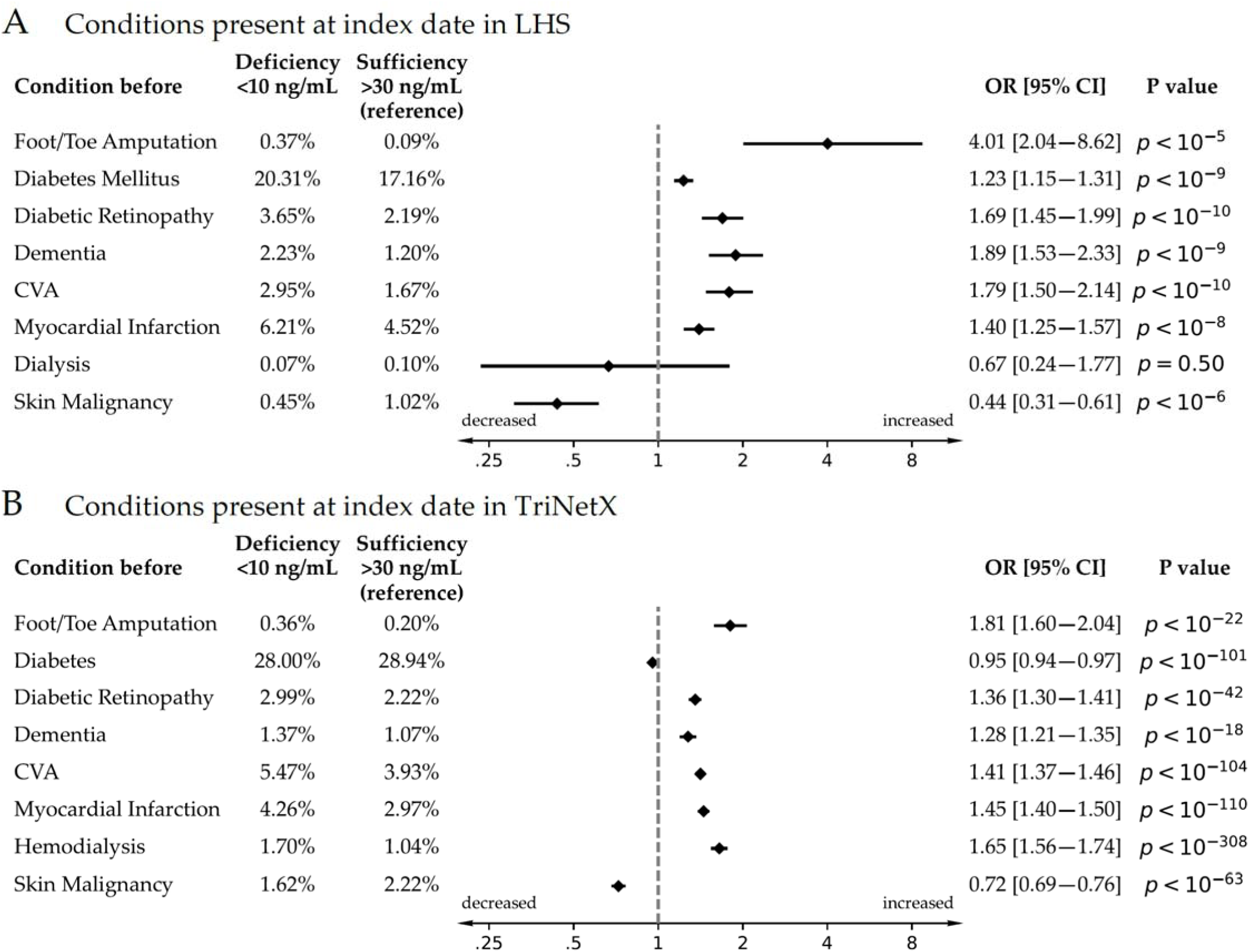
Cross-sectional baseline prevalence of chronic conditions by vitamin D status in LHS (Analysis B) and TriNetX (Analysis C) (A) Odds ratios (OR) and 95% confidence intervals for preexisting conditions at cohort entry in the Leumit Health Services (LHS) matched cohort, comparing individuals with severe vitamin D deficiency (<10 ng/mL) to those with sufficient levels (>30 ng/mL). (B) Corresponding analysis from the TriNetX propensity-matched cohort. Conditions such as diabetes mellitus, diabetic retinopathy, dementia, myocardial infarction, and heart failure were consistently more prevalent in the vitamin D–deficient groups across both datasets. Foot/toe amputations showed the strongest association. In contrast, skin malignancy was significantly less common among individuals with low vitamin D, likely reflecting reduced cumulative UV exposure. These robust and replicated imbalances underscore the biological relevance of vitamin D status beyond demographic and behavioral confounding.

### Analysis C: US TriNetX matched cohort of 25(OH)D deficient (<10ng/mL) vs. sufficient individuals - baseline

To validate these findings, we performed a similar analysis in an independent population using TriNetX, a global federated health research network that aggregates electronic health records from more than 70 healthcare organizations covering over 120 million patients. Eligible members of the US network, with a documented serum 25(OH)D measurement obtained within one year of a recorded BMI value. Individuals with severe deficiency (<10 ng/mL) were compared with those having sufficient levels (>30 ng/mL) using propensity-score matching on age, sex, smoking status, Black or African-American ethnicity, essential hypertension, diabetes mellitus diagnosis, BMI, blood-pressure categories, and hemoglobin A1c categories. Table 2 presents the clinical characteristics of the cohort at baseline, showing well balanced groups and near-identical distributions of age (mean ∼ 54 years), sex (≈ 65 % female), cardiometabolic risk factors, and laboratory measures.

**Table 2.**
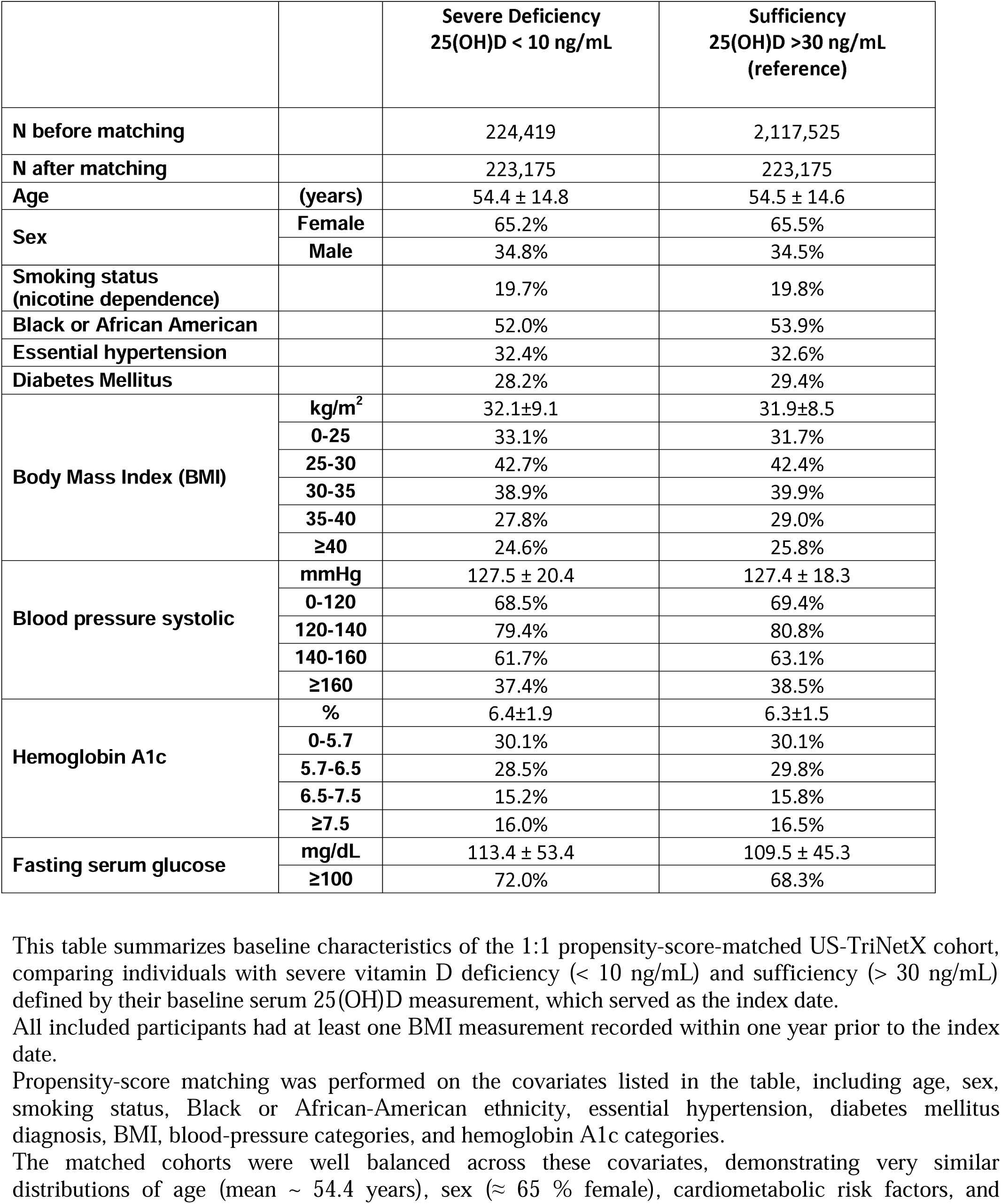
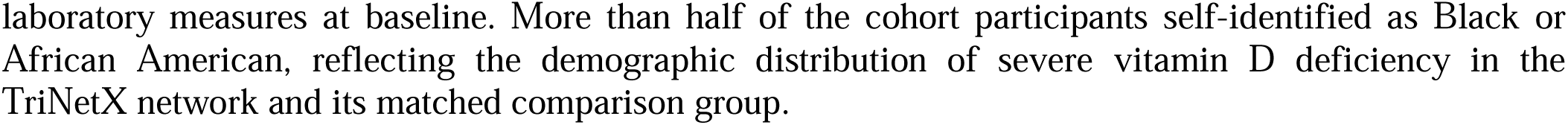
Demographic and clinical characteristics of the US-TriNetX matched cohort at baseline (Analysis C)

|  |  | **Severe Deficiency**<br>**25(OH)D < 10 ng/mL** | **Sufficiency**<br>**25(OH)D >30 ng/mL**<br>**(reference)** |
| --- | --- | --- | --- |
| **N before matching** |  | 224,419 | 2,117,525 |
| **N after matching** |  | 223,175 | 223,175 |
| **Age** | **(years)** | 54.4 ± 14.8 | 54.5 ± 14.6 |
| **Sex** | **Female** | 65.2% | 65.5% |
|  | **Male** | 34.8% | 34.5% |
| **Smoking status**<br>**(nicotine dependence)** |  | 19.7% | 19.8% |
| **Black or African American** |  | 52.0% | 53.9% |
| **Essential hypertension** |  | 32.4% | 32.6% |
| **Diabetes Mellitus** |  | 28.2% | 29.4% |
| **Body Mass Index (BMI)** | **kg/m<sup>2</sup>** | 32.1±9.1 | 31.9±8.5 |
|  | **0-25** | 33.1% | 31.7% |
|  | **25-30** | 42.7% | 42.4% |
|  | **30-35** | 38.9% | 39.9% |
|  | **35-40** | 27.8% | 29.0% |
|  | **≥40** | 24.6% | 25.8% |
| **Blood pressure systolic** | **mmHg** | 127.5 ± 20.4 | 127.4 ± 18.3 |
|  | **0-120** | 68.5% | 69.4% |
|  | **120-140** | 79.4% | 80.8% |
|  | **140-160** | 61.7% | 63.1% |
|  | **≥160** | 37.4% | 38.5% |
| **Hemoglobin A1c** | **%** | 6.4±1.9 | 6.3±1.5 |
|  | **0-5.7** | 30.1% | 30.1% |
|  | **5.7-6.5** | 28.5% | 29.8% |
|  | **6.5-7.5** | 15.2% | 15.8% |
|  | **≥7.5** | 16.0% | 16.5% |
| **Fasting serum glucose** | **mg/dL** | 113.4 ± 53.4 | 109.5 ± 45.3 |
|  | **≥100** | 72.0% | 68.3% |

This table summarizes baseline characteristics of the 1:1 propensity-score-matched US-TriNetX cohort, comparing individuals with severe vitamin D deficiency (< 10 ng/mL) and sufficiency (> 30 ng/mL) defined by their baseline serum 25(OH)D measurement, which served as the index date.
All included participants had at least one BMI measurement recorded within one year prior to the index date.
Propensity-score matching was performed on the covariates listed in the table, including age, sex, smoking status, Black or African-American ethnicity, essential hypertension, diabetes mellitus diagnosis, BMI, blood-pressure categories, and hemoglobin A1c categories.
The matched cohorts were well balanced across these covariates, demonstrating very similar distributions of age (mean ~ 54.4 years), sex (~ 65 % female), cardiometabolic risk factors, and
laboratory measures at baseline. More than half of the cohort participants self-identified as Black or African American, reflecting the demographic distribution of severe vitamin D deficiency in the TriNetX network and its matched comparison group.

**Table 3.**
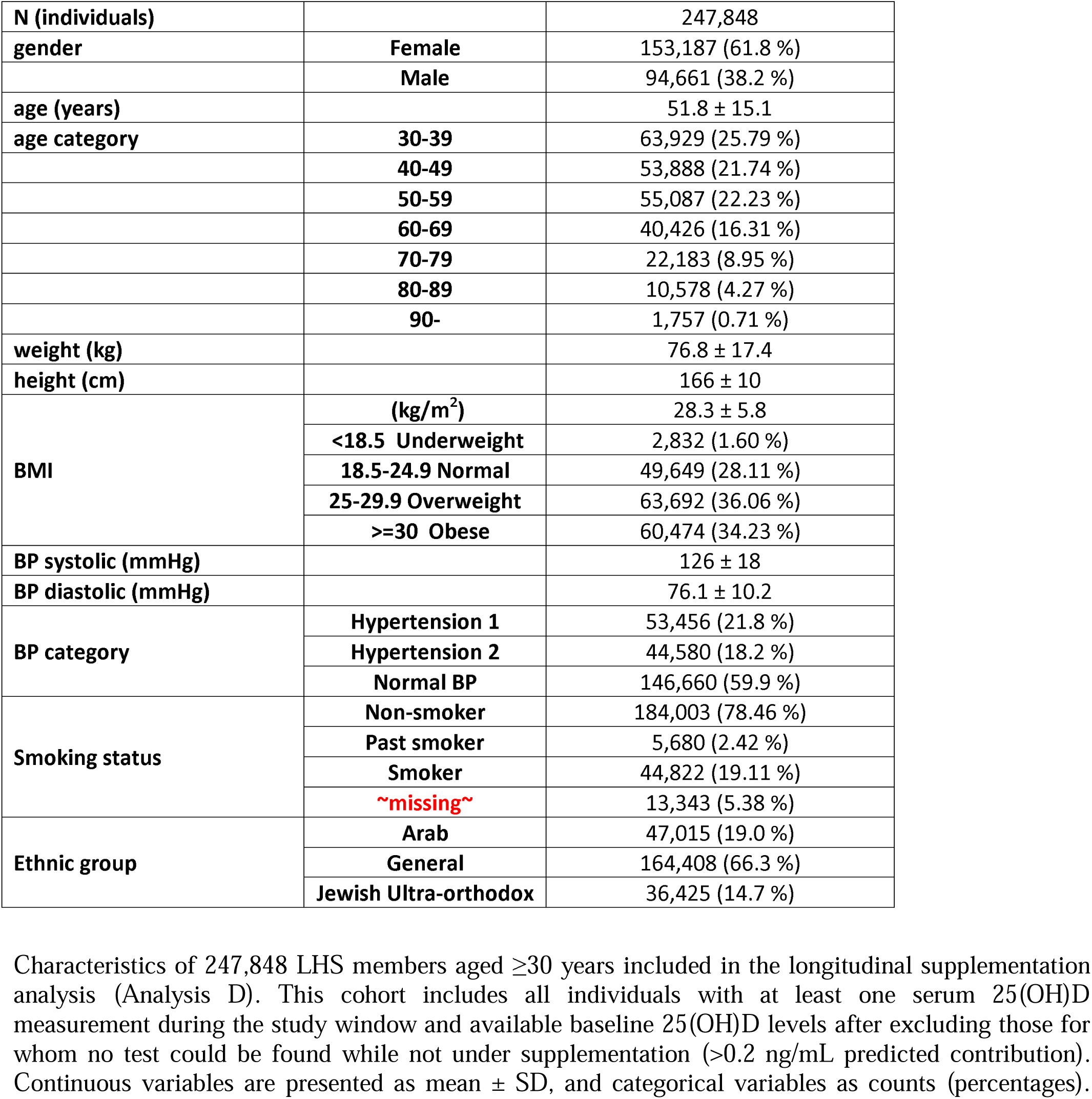
Demographic and clinical characteristics of the Leumit Health Services (LHS) longitudinal cohort used for analysis of vitamin D supplementation (Analysis D)

|  |  |  |
| --- | --- | --- |
| **N (individuals)** |  | 247,848 |
| **gender** | **Female** | 153,187 (61.8 %) |
|  | **Male** | 94,661 (38.2 %) |
| **age (years)** |  | 51.8 ± 15.1 |
| **age category** | **30-39** | 63,929 (25.79 %) |
|  | **40-49** | 53,888 (21.74 %) |
|  | **50-59** | 55,087 (22.23 %) |
|  | **60-69** | 40,426 (16.31 %) |
|  | **70-79** | 22,183 (8.95 %) |
|  | **80-89** | 10,578 (4.27 %) |
|  | **90-** | 1,757 (0.71 %) |
| **weight (kg)** |  | 76.8 ± 17.4 |
| **height (cm)** |  | 166 ± 10 |
| **BMI** | **(kg/m<sup>2</sup>)** | 28.3 ± 5.8 |
|  | **<18.5 Underweight** | 2,832 (1.60 %) |
|  | **18.5-24.9 Normal** | 49,649 (28.11 %) |
|  | **25-29.9 Overweight** | 63,692 (36.06 %) |
|  | **≥30 Obese** | 60,474 (34.23 %) |
| **BP systolic (mmHg)** |  | 126 ± 18 |
| **BP diastolic (mmHg)** |  | 76.1 ± 10.2 |
| **BP category** | **Hypertension 1** | 53,456 (21.8 %) |
|  | **Hypertension 2** | 44,580 (18.2 %) |
|  | **Normal BP** | 146,660 (59.9 %) |
| **Smoking status** | **Non-smoker** | 184,003 (78.46 %) |
|  | **Past smoker** | 5,680 (2.42 %) |
|  | **Smoker** | 44,822 (19.11 %) |
|  | **~missing~** | 13,343 (5.38 %) |
| **Ethnic group** | **Arab** | 47,015 (19.0 %) |
|  | **General** | 164,408 (66.3 %) |
|  | **Jewish Ultra-orthodox** | 36,425 (14.7 %) |

Characteristics of 247,848 LHS members aged ≥30 years included in the longitudinal supplementation analysis (Analysis D). This cohort includes all individuals with at least one serum 25(OH)D measurement during the study window and available baseline 25(OH)D levels after excluding those for whom no test could be found while not under supplementation (>0.2 ng/mL predicted contribution). Continuous variables are presented as mean ± SD, and categorical variables as counts (percentages).

Baseline clinical associations in TriNetX (Figure 3B) were directionally consistent with those in the LHS cohort, though somewhat attenuated, likely because cohort classification in LHS used season-adjusted 25(OH)D levels, whereas TriNetX relied on unadjusted measurements and forced matching for diabetes-related laboratory variables, which may have partially diluted some of the associations.

Nevertheless, the TriNetX cohort showed the same pattern of higher odds of chronic conditions at baseline. Notably, the deficient group had significantly higher rates of diabetes**-**related and cardiovascular complications, including foot/toe amputation (OR 1.81, 95 % CI [1.60–2.04], p = 6.2 × 10 ²¹), diabetic retinopathy (OR 1.36 [1.30–1.41], p < 10□□³), dementia (OR 1.28 [1.21–1.35], p < 10□¹□), CVA (OR 1.41 [1.37–1.46], p< 10□¹²□), myocardial infarction (OR 1.45 [1.40–1.50], p < 10□¹□□), and hemodialysis (OR 1.65 [1.56–1.74], p < 10). In contrast, the odds of skin malignancy were significantly lower in the vitamin D-deficient group (OR 0.72, 95 % CI [0.69–0.76], p = 6.7 × 10□□□), again suggesting lower cumulative ultraviolet skin exposure among individuals with low 25(OH)D levels.

The consistency of these baseline differences across two large and demographically distinct cohorts reinforces the association between severe vitamin D deficiency and a broad profile of chronic disease burden not attributable to age, sex, BMI, smoking status, blood pressure, ethnicity, or other matched covariates.

### Longitudinal impact of 25(OH)D deficiency in the LHS and TriNetX cohorts

To evaluate the longitudinal impact of vitamin D deficiency, we conducted survival analyses excluding individuals who had the outcome of interest at baseline or who developed it within six months of the baseline measurement. Both cohorts were then followed longitudinally for up to 10 years from the index date, applying a six-month safety lag. In LHS, we used Cox proportional-hazards models to estimate adjusted hazard ratios (aHR) in the matched cohort with adjustment for the following covariates: age (continuous), sex, socioeconomic status (fine categories), baseline diabetes (defined by ICD-9 diagnosis or HbA1c >6.5), smoking status (never, past, current, or missing), BMI (fine categories plus an offset from the category lower boundary), kidney function (GFR categories), and albuminuria (urinary albumin/creatinine categories). Figure 4 (left panel) displays the Kaplan–Meier survival curves together with the aHRs, 95% confidence intervals, and p-values for LHS.

**Figure 4:**
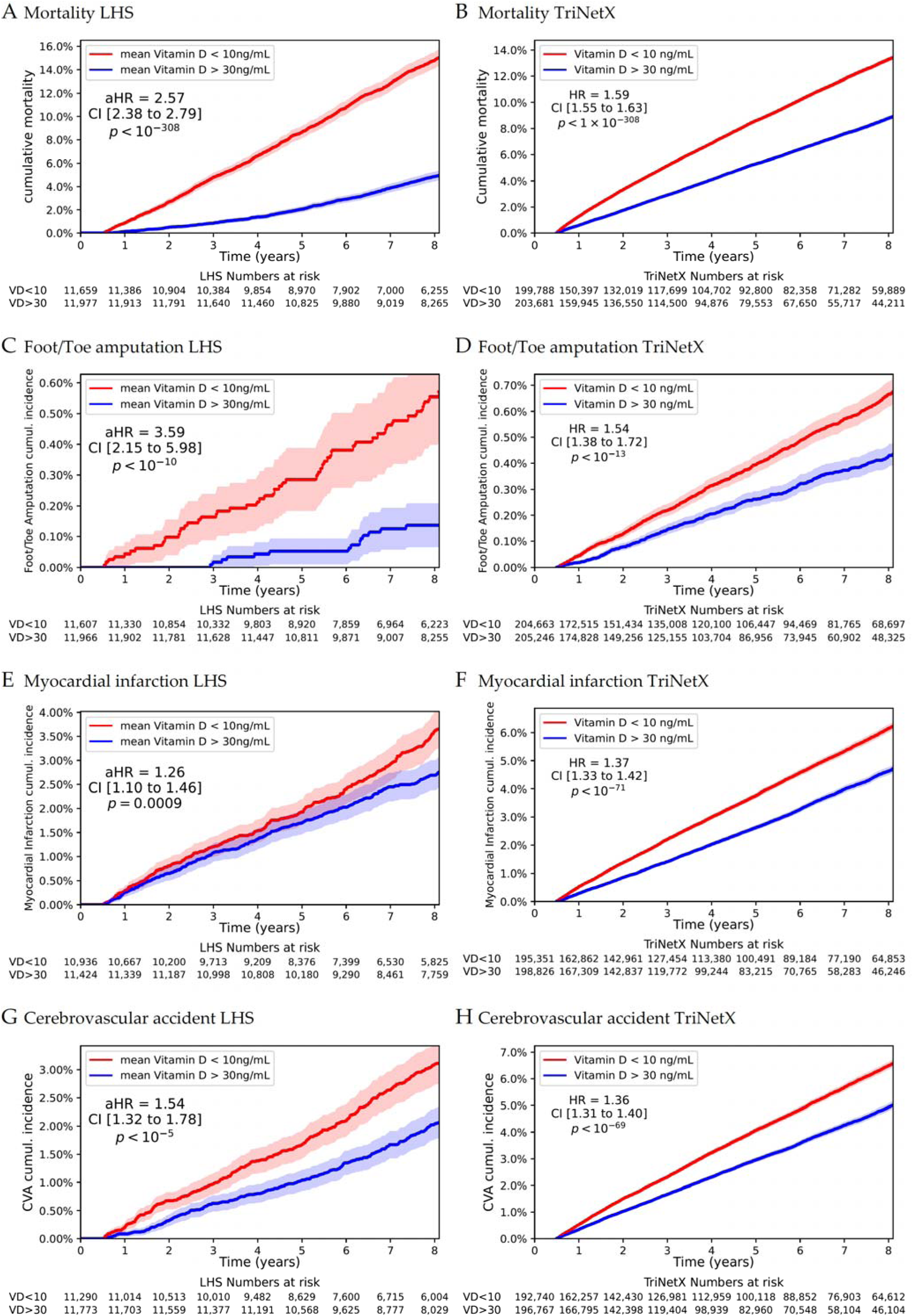
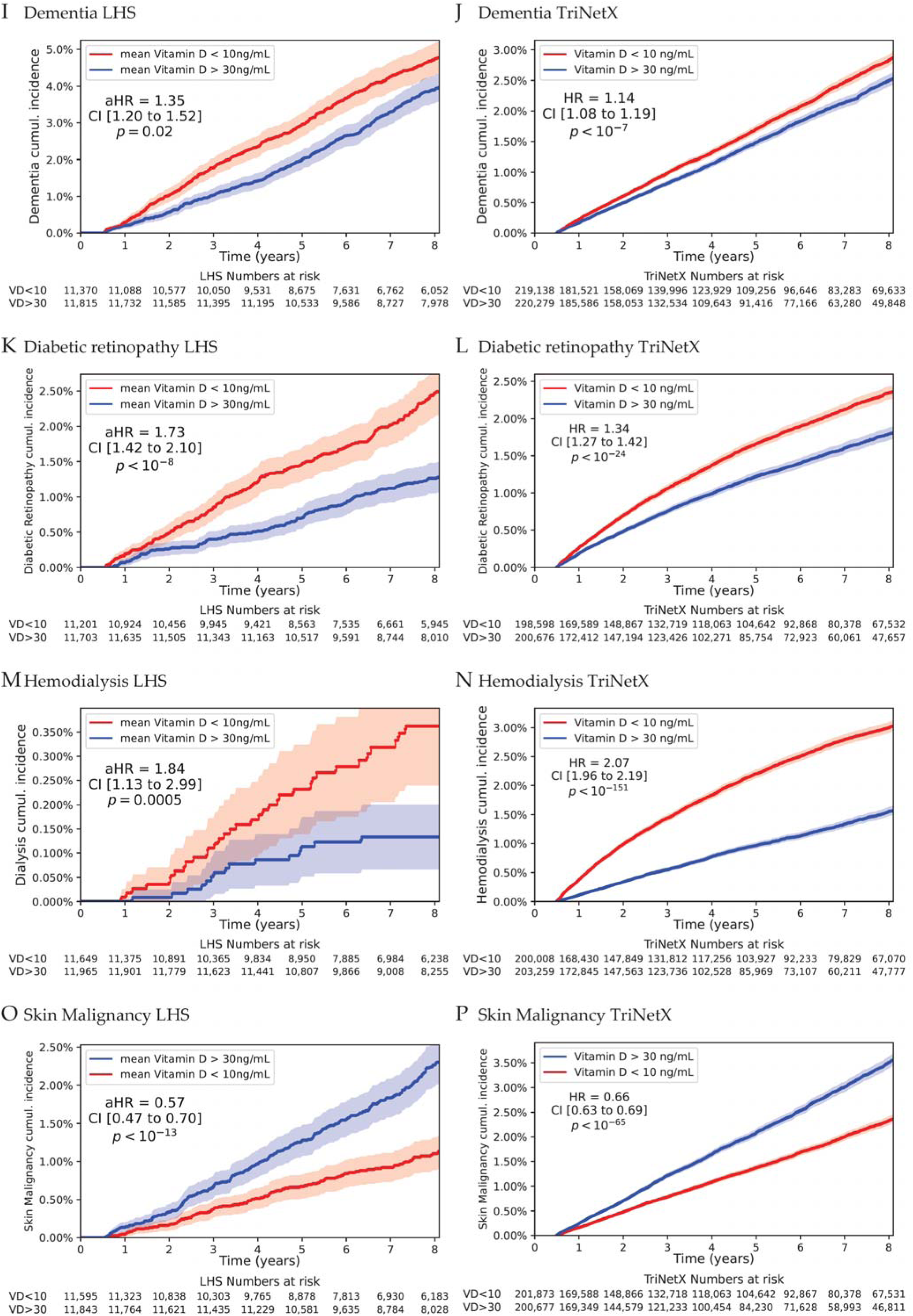
Longitudinal risk of mortality and incident disease by vitamin D status in LHS (left-panel) and TriNetX (right-panel) Kaplan-Meier survival and cumulative incidence curves for matched cohorts of adults ≥30 years, comparing individuals with baseline serum 25(OH)D <10 ng/mL vs >30 ng/mL across two independent cohorts: Leumit Health Services (LHS, left panels) and TriNetX (right panels). Time is years since the index (first qualifying 25(OH)D test). Numbers at risk are shown below each plot. Only incident events occurring ≥6 months after the index date were included; individuals with events in the first year after index were excluded prior to analysis. Panels: (A–B) All-cause mortality; (C–D) Foot/toe amputation; (E–F) Myocardial infarction; (G–H) Cerebrovascular accident; (I–J) Dementia; (K–L) Diabetic Retinopathy; (M–N) Hemodialysis initiation; (O–P) Skin malignancy. LHS (left): Adjusted hazard ratios (aHRs), 95% CI, and P values are from Cox models fit in the 1:1 exact-matched cohort (Table 1) and further adjusted for age (continuous), sex, socioeconomic status (fine categories), baseline diabetes (ICD-9 diagnosis or HbA1c >6.5), smoking status (never, past, current, or missing), BMI (fine categories plus an offset from the category lower boundary), kidney function (GFR categories), and albuminuria (urinary albumin/creatinine categories). The reference category is vitamin D sufficiency (>30 ng/mL). TriNetX (right): Hazard Ratios (HRs), 95% CIs, and P values are from Cox models in 1:1 propensity score matched cohorts (Table 2), balanced on age at index, current age, sex, ethnicity, race, diabetes diagnosis, BMI, GFR, glucose, hemoglobin, AST, ALT, HDL, LDL, and HbA1c. A 6-month lag was applied as above. The reference category is vitamin D sufficiency (>30 ng/mL).

We performed analogous analyses in TriNetX, using Cox proportional-hazards models to estimate hazard ratios (HRs) for the same outcomes (Figure 4, right panel).

This analysis revealed a consistently higher cumulative incidence of mortality and major outcomes during follow-up among individuals with severe vitamin D deficiency in both cohorts.

#### LHS cohort

- All-cause mortality: cumulative 8-year incidence ≈ 14 % in the deficient group vs ≈ 8 % in controls; aHR = 2.57 (95 % CI [2.38–2.79], p < 10□³□□)
- Foot/Toe amputation: aHR = 3.59 ([2.15–5.98], p < 10□¹□)
- Myocardial infarction: aHR = 1.26 ([1.10–1.46], p = 0.0009)
- Cerebrovascular accident: aHR = 1.54 ([1.32–1.78], p < 10□□)
- Dementia: aHR = 1.35 ([1.20–1.52], p = 0.02)
- Diabetic retinopathy: aHR = 1.73 ([1.42–2.10], p < 10□□)
- Dialysis: aHR = 1.84 ([1.13–2.99], p = 0.0005)
- Skin malignancy: consistently lower in the deficient group (aHR = 0.57 [0.47–0.70], p < 10□¹³)

#### TriNetX cohort

- All-cause mortality: HR = 1.59 (95 % CI [1.55–1.63], p < 10□³□□)
- Foot/toe amputation: HR = 1.54 ([1.38–1.72], p < 10□¹³)
- Myocardial infarction: HR = 1.37 ([1.33–1.42], p < 10□□¹)
- Cerebrovascular accident: HR = 1.36 ([1.31–1.40], p < 10□□□)
- Dementia: HR = 1.14 ([1.08–1.19], p < 10□□)
- Diabetic retinopathy: HR = 1.34 ([1.27–1.42], p < 10□²□)
- Hemodialysis: HR = 2.07 ([1.96–2.19], p < 10□¹□¹)
- Skin malignancy: significantly lower in the deficient group, HR = 0.66 ([0.63–0.69], p < 10□□□)

In both cohorts, the divergence in risk between the deficient and sufficient groups increased over time, even after excluding events that occurred prior to the index date or during the initial six-month lag period. These associations remained materially unchanged when applying a 12-month lag period to further minimize reverse causation (Supplementary Figure 2).

The replication of these trends in matched cohorts across two large, geographically and structurally distinct healthcare systems reinforces the robustness and generalizability of the observed associations. The consistency in magnitude, direction, and timing of risk divergence across datasets supports a potential contributory role of severe vitamin D deficiency in chronic disease progression and mortality.

### Modeling vitamin D supplementation impact on 25(OH)D levels

To evaluate whether vitamin D represents a modifiable risk factor, we investigated the effect of supplementation on 25(OH)D levels and downstream clinical outcomes. We began by decomposing the variation in serum 25(OH)D levels into two components: one attributable to largely non-modifiable demographic and structural factors—such as BMI, sex, ethnicity, and socioeconomic status—and another potentially modifiable through supplementation. Using a multivariable linear regression model, we predicted log-transformed 25(OH)D levels based on sex, ethnic group, socioeconomic status, BMI category, age, and seasonal harmonics. This model explained 25.4% of the variance in 25(OH)D levels (R² = 0.254) in the population (Supplementary Table 2).

To isolate the contribution of vitamin D supplementation, we quantified the total intake in international units (IU) from all vitamin D products purchased through LHS pharmacies during each of the 4 months preceding a 25(OH)D test, as well as over quarterly and yearly periods. We then modeled the difference between the observed serum 25(OH)D levels and those predicted based on demographic characteristics and BMI, regressing this residual on estimated supplementation amounts, aggregated into predefined time-period bins. Supplement use in the months prior to testing had the strongest effect, with diminishing impact for more distant time windows (Supplementary Table 3). Together, supplementation variables explained an additional 9.% of the variance in 25(OH)D levels not captured by baseline characteristics, demonstrating a quantifiable effect of supplement use on serum 25(OH)D levels.

### Analysis D: Assessing the independent effect of vitamin D supplementation

To assess how vitamin D supplementation influences clinical outcomes, we followed 260,562 Leumit Health Services (LHS) members aged ≥30 years with at least one serum 25(OH)D measurement, representing 84.4% of members in this age range. We excluded individuals for whom no serum 25(OH)D test could be found while not under supplementation (>2 ng/mL predicted contribution), yielding 247,848 participants with baseline 25(OH)D levels available for analysis. For this cohort, we computed two variables:

- **Calendar-adjusted baseline serum 25(OH)D concentration**, categorized as <5, 5–10, 10–15, 15–20, and >20 ng/mL (reference group); and
- **Vitamin D supplementation**, modeled as a time-dependent covariate reflecting the predicted monthly increase in serum 25(OH)D based on pharmacy purchase data, categorized as <0.2 ng/mL (reference), 0.2–5 ng/mL, 5–10 ng/mL, and >10 ng/mL serum-equivalent.

We fitted joint multivariable Cox proportional-hazards models including both baseline 25(OH)D categories and vitamin D supplementation as a time-dependent covariate to estimate associations with all-cause mortality and major clinical outcomes. Models were adjusted for age, sex, smoking status, socioeconomic status, BMI (fine categories plus offset), blood pressure, estimated GFR, albuminuria category, baseline diabetes, and hypertension. Results are shown in Figure 5.

**Figure 5:**
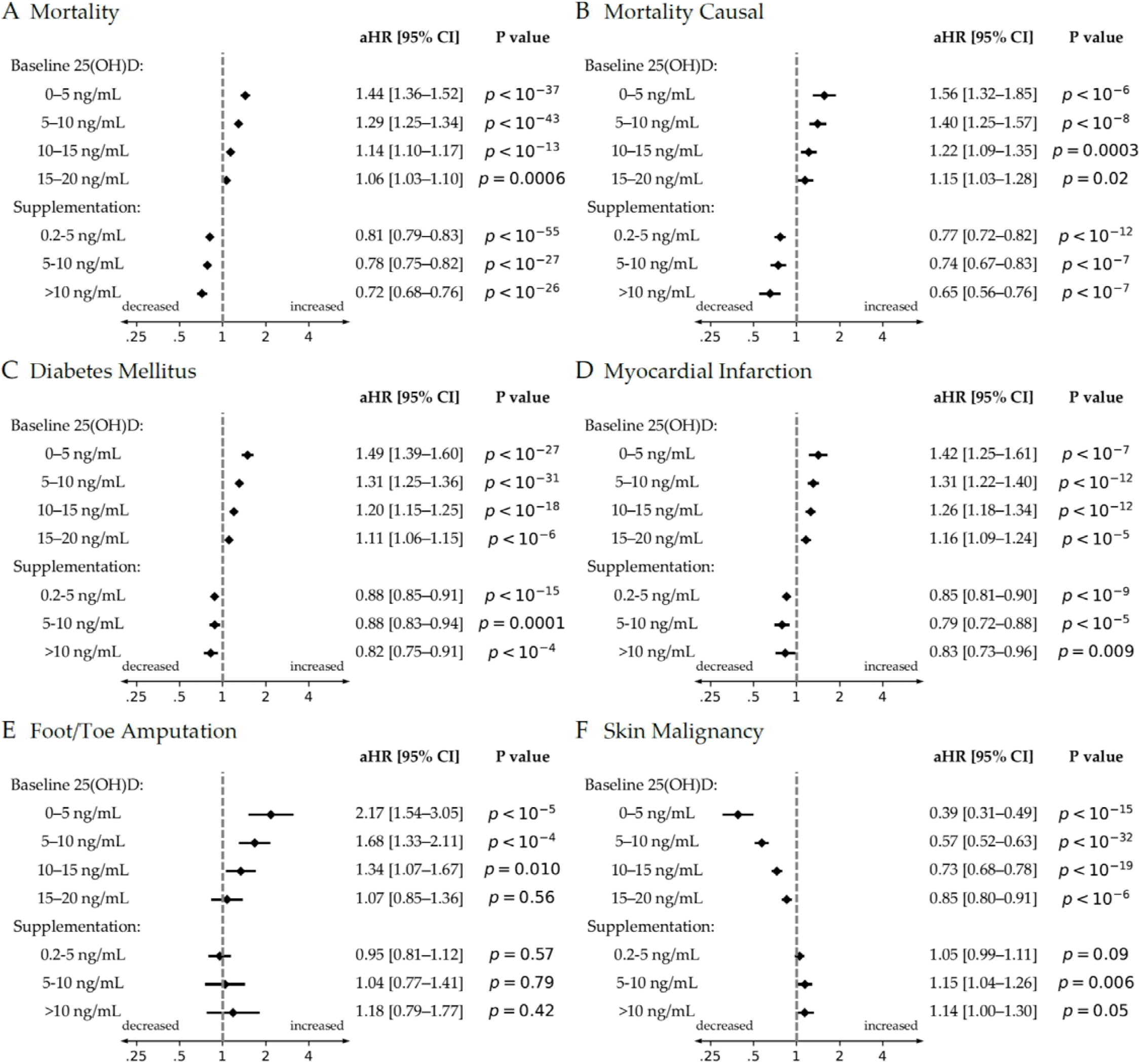
Longitudinal joint effects of baseline vitamin D status and supplementation on health outcomes in LHS (Analysis D) in LHS Adjusted hazard ratios (HRs) with 95% confidence intervals (log scale) are shown for mortality and five clinical outcomes among all Leumit Health Services (LHS) members aged >=30 years who had at least one serum 25(OH)D measurement since 2009. The index date was defined as the first available measurement within the study window. Cox proportional-hazards models jointly included baseline serum 25(OH)D categories (0-5, 5-10, 10-15, 15-20 ng/mL; reference >20 ng/mL) and vitamin D supplementation categories modeled time-dependently from pharmacy dispensing records (0.2-5, 5-10, >10 ng/mL serum-equivalent; reference <0.2 ng/mL). Models were adjusted for age, sex, smoking status, socioeconomic status, BMI (fine categories plus offset), estimated GFR category, albuminuria category, and baseline diabetes, and applied a 6-month lag to minimize reverse causation from early events. Panel B presents results from inverse-probability of treatment stabilized weighting (IPTW) marginal structural Cox models for mortality, using cumulative stabilized weights to account for time-varying confounding of supplementation by prior measured covariates. Stabilized weights were truncated at the 1st and 99th percentiles to reduce the influence of extreme values. This causal-framework sensitivity analysis was performed for all-cause mortality, the primary endpoint. An HR <1 indicates lower risk; HR >1 indicates higher risk. The vertical dashed line marks HR = 1. Panels: (A) Mortality (standard adjusted model), (B) Mortality (IPTW marginal structural model), (C) Diabetes mellitus, (D) Myocardial infarction, (E) Foot/Toe amputation, (F) Skin malignancy. Higher baseline 25(OH)D and increasing supplementation dose were consistently associated with lower risks of outcomes in panels A-D. Panel F served as a negative-control outcome: it showed an inverse association with baseline vitamin D, likely reflecting higher sun exposure among individuals with higher baseline 25(OH)D, but no consistent protective association with supplementation calculated from pharmacy dispensing data.

For mortality (Figure 5A), baseline vitamin D deficiency was associated with a graded, dose–dependent increase in risk: HR = 1.44 [1.36–1.52] for <5 ng/mL; HR = 1.29 [1.25–1.34] for 5–10 ng/mL; HR = 1.14 [1.10–1.17] for 10–15 ng/mL; and HR = 1.06 [1.03–1.10] for 15–20 ng/mL (reference >20 ng/mL; all p < 0.001). Vitamin D supplementation was independently protective in a dose–dependent fashion: HR = 0.72 [0.68–0.76] for >10 ng/mL serum–equivalent; HR = 0.78 [0.75–0.82] for 5–10 ng/mL; and HR = 0.81 [0.79–0.83] for 0.2–5 ng/mL (all p < 10^-26^).

To strengthen causal interpretation, we additionally performed inverse–probability of treatment weighted (IPTW) marginal structural Cox analyses for mortality (Figure 5B). In these causal–framework analyses, the graded association between baseline vitamin D deficiency and mortality remained evident and became slightly stronger: HR = 1.56 [1.32–1.85] for <5 ng/mL; HR = 1.40 [1.25–1.57] for 5–10 ng/mL; HR = 1.22 [1.09–1.35] for 10–15 ng/mL; and HR = 1.15 [1.03–1.28] for 15–20 ng/mL. Supplementation also remained independently protective in a dose–dependent manner, with HR = 0.65 [0.56–0.76] for >10 ng/mL serum–equivalent; HR = 0.74 [0.67–0.83] for 5–10 ng/mL; and HR = 0.77 [0.72–0.82] for 0.2–5 ng/mL (all p < 10^-7^).

For diabetes mellitus (Figure 5C), lower baseline vitamin D was strongly associated with higher risk: HR = 1.49 [1.39–1.60] for <5 ng/mL; HR = 1.31 [1.25–1.36] for 5–10 ng/mL; HR = 1.20 [1.15–1.25] for 10–15 ng/mL; and HR = 1.11 [1.06–1.15] for 15–20 ng/mL (all p < 10^-6^).

Supplementation was again protective: HR = 0.88 [0.85–0.91] for 0.2–5 ng/mL; HR = 0.88 [0.83–0.94] for 5–10 ng/mL; and HR = 0.82 [0.75–0.91] for >10 ng/mL (all p < 0.001).

For myocardial infarction (Figure 5D), low baseline vitamin D was linked to higher risk (HR = 1.42 [1.25–1.61] for <5 ng/mL; HR = 1.31 [1.22–1.40] for 5–10 ng/mL; HR = 1.26 [1.18–1.34] for 10–15 ng/mL; HR = 1.16 [1.09–1.24] for 15–20 ng/mL; all p < 10^-5^), and supplementation again showed significant protection, including HR = 0.85 [0.81–0.90] for 0.2–5 ng/mL, HR = 0.79 [0.72–0.88] for 5–10 ng/mL, and HR = 0.83 [0.73–0.96] for >10 ng/mL.

For foot/toe amputation (Figure 5E), a strong baseline dose–response was evident, with HR = 2.17 [1.54–3.05] for <5 ng/mL; HR = 1.68 [1.33–2.11] for 5–10 ng/mL; and HR = 1.34 [1.07–1.67] for 10–15 ng/mL. No significant protective effect of supplementation was detected, likely due to the limited number of amputation events under supplementation.

For skin malignancy (Figure 5F), baseline vitamin D levels showed an inverse association, consistent with lower UV exposure in vitamin D-deficient individuals, whereas supplementation showed no consistent protective effect (HRs near 1.0). This discordant pattern is particularly informative because it argues against non-specific healthy-user or ultraviolet-exposure confounding as the primary explanation for the supplementation findings.

These findings demonstrate that lower baseline vitamin D levels are associated with increased risks across multiple aging-related outcomes, while supplementation, modeled independently of baseline status, was associated with dose-dependent reductions in mortality, metabolic disease, and cardiovascular risk. Importantly, these associations remained robust in causal-framework sensitivity analyses using inverse-probability weighted marginal structural Cox models, which were specifically designed to reduce bias from time-varying treatment selection and measured confounding.

The consistency of the findings across conventional adjusted models, time-dependent supplementation analyses, and IPTW marginal structural models supports the interpretation that at least part of the observed associations may reflect a modifiable and potentially causal role of vitamin D status in chronic disease prevention and healthy aging, rather than solely residual confounding or reverse causation.

### Risk Reduction and Number Needed to Treat

To evaluate the clinical utility of vitamin D supplementation, we estimated the 10-year number needed to treat (NNT) to prevent one all-cause mortality event, stratified by age group (≥30, ≥50, and ≥70 years), baseline serum 25(OH)D category, and the predicted supplementation-related increase in serum levels (Figure 6). Because mortality represents the only outcome not subject to potential competing-risk bias, NNT estimation was restricted to all-cause mortality.

**Figure 6:**
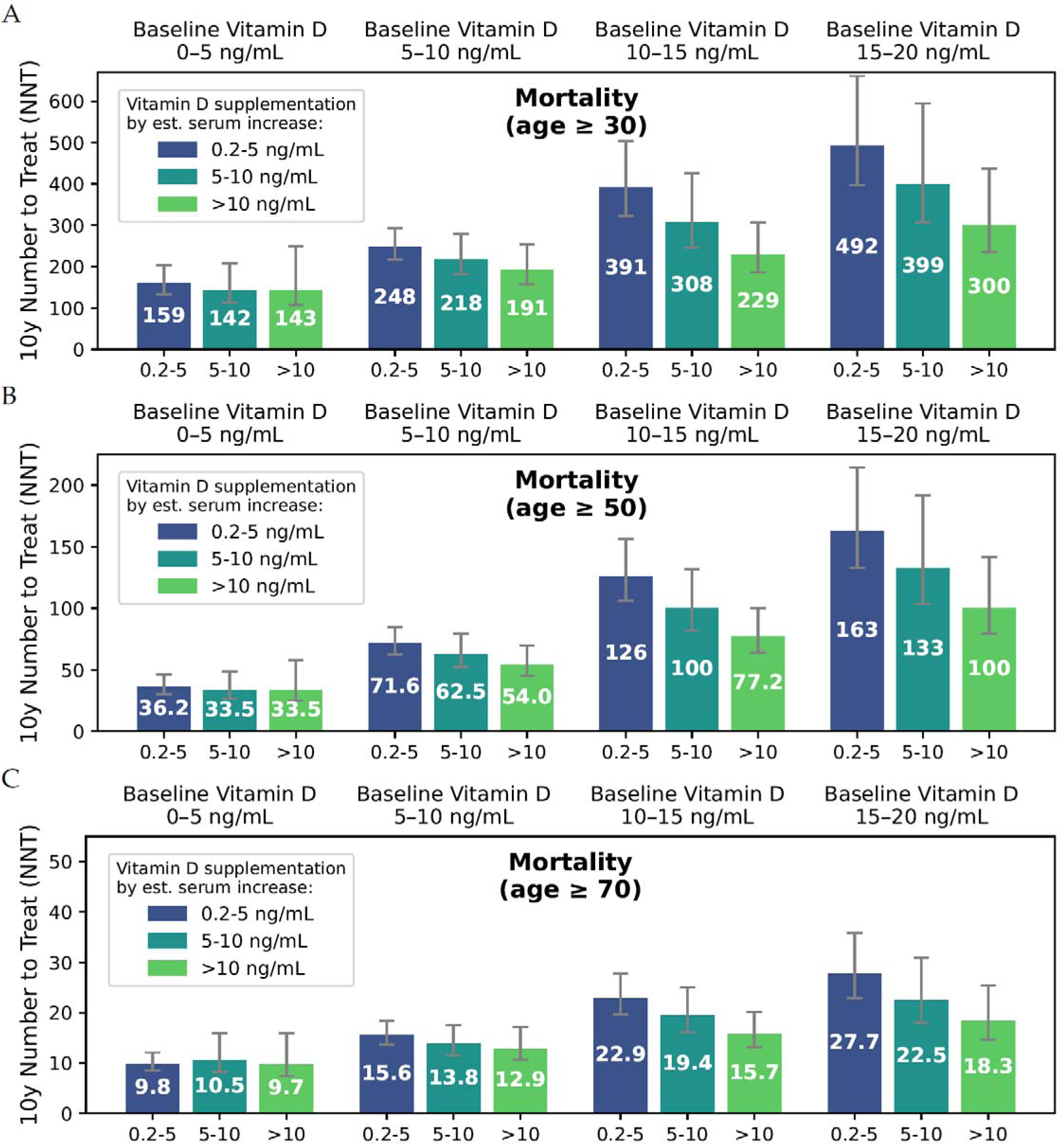
Estimated 10-year number needed to treat (NNT) for prevention of all-cause mortality by age, baseline vitamin D status, and supplementation level in LHS (Analysis D). Panels A–C show the estimated 10-year number needed to treat (NNT) to prevent one death for each vitamin D supplementation category (color-coded bars), stratified by baseline serum 25(OH)D concentration (groups of columns) and by age threshold (rows: ≥30, ≥50, ≥70 years). Supplementation categories are defined by the modeled increase in serum 25(OH)D (ng/mL). Error bars represent 95 % confidence intervals. NNT values were derived from absolute risk reductions estimated from the joint time-dependent Cox proportional-hazards models (see Methods). NNT declined markedly with older age and with lower baseline 25(OH)D (<10 ng/mL), reaching approximately 10–15 in older adults with very low baseline levels—indicating a substantial preventive benefit of vitamin D supplementation in these high-risk groups.

Protective effects were evident at all ages and for all baseline vitamin D categories below 20 ng/mL, with NNTs declining sharply with advancing age. Among participants with baseline 25(OH)D < 5 ng/mL, NNTs at age 30 ranged from 142–159 depending on supplementation dose, but improved markedly to 33.5–36.2 at age 50 and as low as 9.7–10.5 at age 70. For those starting at 5–10 ng/mL, NNTs similarly decreased from 191–248 at age 30 to 54.0–71.6 at age 50 and 12.9–15.6 at age 70. At baseline levels of 10–15 ng/mL, NNTs dropped from 229–391 at age 30 to 77.2–126 at age 50 and 15.7–22.9 at age 70. Even among individuals with baseline 15–20 ng/mL, NNTs remained clinically meaningful in older adults, declining from 300–492 at age 30 to 100–163 at age 50 and 18.3–27.7 at age 70.

This pronounced age-gradient reflects the rising absolute mortality risk in older adults and highlights the potentially greater absolute benefit of vitamin D supplementation in individuals with severe or moderate deficiency, especially at age 70 years and above. Collectively, these results show that vitamin D supplementation could yield substantial clinical benefit, especially for those with baseline 25(OH)D < 10 ng/mL, by preventing mortality events at relatively low NNTs in older populations.

## Discussion

In this dual-cohort longitudinal analysis leveraging real-world data from over 120 million individuals across two countries, severe vitamin D deficiency consistently identified a systemic state of increased aging-related vulnerability spanning metabolic, vascular, renal, neurodegenerative, and mortality-related domains. Importantly, our analyses suggest that vitamin D supplementation, modeled through pharmacy records and validated serum responses, is associated with significant reductions in risk across a wide range of clinical outcomes. The protective associations were consistent, dose-dependent, and remained significant even after controlling for confounding demographic, behavioral, and socioeconomic factors. Notably, the observed associations extended across multiple major age-related outcomes and organ systems, supporting the concept that vitamin D status may function as a broad determinant of systemic aging vulnerability and multimorbidity accumulation rather than merely a disease-specific biomarker.

The LHS matched cohort design allowed for tight baseline matching of demographic and clinical risk factors, including BMI, blood pressure, diabetes, smoking status, and kidney disease. Findings were replicated in an independent, US-based federated health network (TriNetX), enhancing external validity. A major strength of this study is the integration of detailed laboratory, demographic, and pharmacy data from a national health organization, enabling precise classification of baseline vitamin D status and estimation of serum responses to supplementation. We leveraged these data to construct a time-dependent covariate for vitamin D supplementation, derived from pharmacy-dispensed records and calibrated to predict serum 25(OH)D changes over time. This approach helped distinguish the potentially modifiable effect of supplementation from the confounding influence of demographic, socioeconomic, and obesity-related determinants of vitamin D deficiency. Importantly, supplementation exposure was modeled longitudinally throughout follow-up, enabling assessment of dynamic exposure-response relationships rather than reliance on baseline supplementation status alone.

The supplementation covariate was based on the residual variation in 25(OH)D unexplained by demographic and clinical predictors (age, sex, BMI, ethnicity, socioeconomic status, and season). As a result, subgroup differences in these baseline determinants were inherently accounted for, while the same variables were also included as covariates in the multivariable Cox models. This design minimizes bias from population heterogeneity and provides a robust framework for assessing the independent, time-updated effect of vitamin D supplementation on outcomes. To further strengthen causal interpretation, we additionally implemented inverse-probability of treatment weighted (IPTW) marginal structural Cox models for all-cause mortality. These models were specifically designed to address time-varying treatment selection and measured confounding in longitudinal supplementation analyses. Importantly, the IPTW analyses yielded results that were directionally consistent with, and in several cases stronger than, the conventional adjusted Cox models, supporting the robustness of the observed supplementation associations. The absence of a corresponding protective association for skin malignancy, despite strong baseline vitamin D associations likely driven by sun exposure, further argues against simple healthy-user or UV-exposure confounding as the sole explanation for the supplementation findings.

Our findings revealed strikingly low numbers needed to treat (NNTs) for all-cause mortality. Notably, the estimated NNTs in older severely deficient adults fall within ranges considered clinically meaningful for established preventive interventions. Even moderate supplementation (predicted serum increase of 10–20 ng/mL) in this group yielded substantial absolute benefit. Importantly, benefit extended beyond the severely deficient: individuals with baseline 25(OH)D of 15–20 ng/mL also showed meaningful reductions in mortality risk, with NNTs of roughly 18–28 depending on supplementation level. These results highlight vitamin D supplementation as a scalable, low-cost intervention with significant public-health potential when targeted to individuals with suboptimal baseline levels. From a geroscience perspective, these findings are notable because interventions capable of simultaneously influencing mortality, cardiometabolic disease, vascular complications, and functional decline remain relatively uncommon in population-scale human datasets.

Previous randomized controlled trials (RCTs) of vitamin D supplementation have yielded inconsistent results for chronic disease and mortality outcomes (Zittermann et al. 2017; Davidson et al. 2013; Chen et al. 2024; Zhang et al. 2019). Several factors likely explain this discrepancy. Most RCTs enrolled participants regardless of baseline vitamin D status, resulting in limited power to detect benefits confined to the severely deficient subgroups, where our strongest associations were observed, particularly among older adults. In addition, background supplement use in placebo groups and incomplete adherence diluted between-group differences in achieved 25(OH)D levels, leading to underestimation of true effects. Moreover, many trials used fixed low doses, without achieving the serum repletion levels associated with reduced mortality and morbidity in our real-world data. These limitations help reconcile the apparent divergence between large pragmatic RCTs and the consistent, dose-dependent associations observed in our cohorts.

The biological mechanisms underlying these protective effects remain to be fully elucidated. One plausible explanation is that vitamin D status influences shared upstream aging-related pathways, potentially including chronic inflammation, immune dysregulation, endothelial dysfunction, microbial-host interactions, and impaired tissue resilience (Schwalfenberg 2011; Merzon et al. 2020; Israel et al. 2022; Lagishetty et al. 2011; Singh et al. 2020). The breadth of associated outcomes observed across cardiovascular, metabolic, renal, neurodegenerative, and mortality-related domains is compatible with mechanisms operating at the level of systemic organismal resilience rather than isolated disease-specific pathways. However, the precise biological mechanisms underlying these associations remain incompletely understood and require further mechanistic investigation. Supporting this hypothesis is the breadth of conditions impacted—including diabetes, cardiovascular disease, and all-cause mortality—which may share common microbial or immune-mediated origins. A particularly notable observation was the significant reduction in foot and toe amputations among supplemented individuals. Given that such amputations are frequently precipitated by persistent infection, this finding further supports a microbiologically mediated mechanism (Joshi et al. 2023). Our findings raise the possibility that severe vitamin D deficiency may contribute to the elevated burden of lower-extremity complications observed in certain high-risk population subgroups. More broadly, the simultaneous associations observed across cardiovascular, metabolic, renal, neurologic, and mortality endpoints are compatible with the possibility that vitamin D status influences shared upstream aging-related pathways, potentially including chronic inflammation, immune dysregulation, microbial-host interactions, endothelial dysfunction, and impaired tissue resilience.

This study has limitations. As an observational analysis, residual confounding cannot be entirely excluded despite extensive matching, adjustment, and validation across two independent cohorts. Vitamin D supplementation may also co-occur with other unmeasured health-promoting behaviors. In addition, our exposure definition relied on pharmacy-dispensed purchases within LHS–affiliated pharmacies and may not fully capture over-the-counter acquisitions through untracked channels. Nevertheless, the strength, consistency, and dose-dependent pattern of associations across cohorts support a genuine underlying relationship. Importantly, the persistence of the associations within IPTW marginal structural models suggests that the observed protective effects are unlikely to be explained solely by measured treatment-selection bias or time-varying confounding. Although standard Cox models treat death as a censoring event, mortality itself functions as a competing outcome for nonfatal events. Finally, although causal inference from observational data necessarily depends on assumptions of exchangeability and adequate capture of confounders, the convergence of findings across conventional multivariable models, longitudinal time-dependent supplementation analyses, dose-response relationships, and IPTW causal-framework analyses substantially strengthens the plausibility of a modifiable biological effect.

In conclusion, our findings support severe vitamin D deficiency as a prevalent and potentially modifiable determinant of aging-related vulnerability across multiple organ systems. The consistency of the associations across independent cohorts, longitudinal exposure modeling, dose-response relationships, and causal-framework sensitivity analyses supports the plausibility that vitamin D status may influence systemic resilience and multimorbidity trajectories during aging. These findings provide a population-scale framework for future mechanistic and interventional studies investigating vitamin D biology within the context of human aging.

## Data Availability

Access to the LHS data used in this study are restricted to researchers approved by the IRB.
The TriNetX data used in this study is limited to members of the TriNetX network.

## Author contributions

Conceptualization: AI, EMe, EMa

Methodology: AI, EMe

Investigation: AI, SI, EMe

Writing – original draft: AI

Writing – review & editing: AI, AW, SI, SA, SV, EMa, EMe

## Materials & Correspondence

Correspondence and requests for materials should be addressed to AI

## Abbreviations

aOR: adjusted Odds Ratio
BMI: Body Mass Index
BP: Blood pressure
CNS: Central nervous system
EHR: Electronic health records
HR: Hazard Ratio
ICD-9: International Classification of Diseases, Ninth Revision
LHS: Leumit Health Services
OR: Odds Ratio PO: Per os
PSM: Propensity score matching
SES: Socioeconomic status
SMD: Standardized Mean Difference

