## Supplementary Tables and Figures for "Vitamin D Deficiency, Supplementation, and Aging-Related Morbidity and Mortality Across Two Longitudinal Population Cohorts"

### Supplementary Table 1. Seasonal Regression Model of Log-Transformed Serum Vitamin D Levels

| **Variable** | **Coefficient (β)** | **Std. Error** | **t-Statistic** | **P-value** | **95% CI Lower** | **95% CI Upper** |
| --- | --- | --- | --- | --- | --- | --- |
| Intercept | 2.8958 | 0.0005 | 5626.8609 | <0.0001 | 2.8948 | 2.8968 |
| season_sin | -0.1008 | 0.0007 | -135.9558 | <0.0001 | -0.1022 | -0.0993 |
| season_cos | -0.1088 | 0.0007 | -152.4612 | <0.0001 | -0.1102 | -0.1074 |
| season_sin² | -0.0160 | 0.0007 | -22.5060 | <0.0001 | -0.0174 | -0.0146 |
| season_cos² | 0.0068 | 0.0007 | 9.1851 | <0.0001 | 0.0054 | 0.0083 |

**Model Summary:**

- Dependent variable: log_vitamin_d
- R-squared = 0.040
- Adjusted R-squared = 0.040
- F-statistic = 10,473 (p < 0.0001)

This regression model estimates the seasonal variation in serum 25(OH)D levels using sinusoidal and second-harmonic terms (sine and cosine). The outcome variable was the natural logarithm of vitamin D levels. Seasonal terms are based on calendar date and are independent of individual-level characteristics. The analysis includes over 1.65 million observations from Leumit Health Services (LHS). The model explains a modest portion of variance (R^2^ = 0.040), reflecting significant impact of seasonality, but also high interindividual variability in serum 25(OH)D.

### Supplementary Table 2. Multivariable OLS Regression Predicting Log-Transformed Serum Vitamin D Levels

| **Variable** | **Coefficient** | **Std. Error** | **t-Statistic** | **P-value** | **95% CI Lower** | **95% CI Upper** |
| --- | --- | --- | --- | --- | --- | --- |
| Intercept | 2.7542 | 0.0033 | 834.3379 | <0.0001 | 2.7477 | 2.7606 |
| Female, Arab | -0.5713 | 0.0020 | -291.9690 | <0.0001 | -0.5752 | -0.5675 |
| Female, Ultra-Orthodox | -0.1533 | 0.0019 | -79.4336 | <0.0001 | -0.1571 | -0.1495 |
| Male, General | 0.0181 | 0.0014 | 12.8078 | <0.0001 | 0.0153 | 0.0209 |
| Male, Arab | -0.1802 | 0.0026 | -70.6107 | <0.0001 | -0.1852 | -0.1752 |
| Male, Ultra-Orthodox | -0.1031 | 0.0025 | -40.6715 | <0.0001 | -0.1081 | -0.0982 |
| SES 4–6 | 0.0788 | 0.0025 | 32.0119 | <0.0001 | 0.0739 | 0.0836 |
| SES 7–9 | 0.0841 | 0.0026 | 32.3250 | <0.0001 | 0.0790 | 0.0892 |
| SES 10–11 | 0.1302 | 0.0027 | 47.7027 | <0.0001 | 0.1249 | 0.1356 |
| SES 12–14 | 0.1316 | 0.0028 | 46.5542 | <0.0001 | 0.1260 | 0.1371 |
| SES ≥15 | 0.1437 | 0.0032 | 44.6216 | <0.0001 | 0.1374 | 0.1500 |
| SES Missing | 0.1180 | 0.0031 | 38.3047 | <0.0001 | 0.1119 | 0.1240 |
| BMI <18.5 (Underweight) | -0.0321 | 0.0048 | -6.6179 | <0.0001 | -0.0416 | -0.0226 |
| BMI 25–29.9 (Overweight) | -0.0657 | 0.0013 | -48.7713 | <0.0001 | -0.0684 | -0.0631 |
| BMI 30–34.9 (Obese I) | -0.1231 | 0.0016 | -76.8983 | <0.0001 | -0.1262 | -0.1199 |
| BMI 35–39.9 (Obese II) | -0.1794 | 0.0022 | -81.6797 | <0.0001 | -0.1837 | -0.1751 |
| BMI ≥40 (Obese III) | -0.2221 | 0.0028 | -79.4461 | <0.0001 | -0.2276 | -0.2166 |
| BMI per unit above boundary | -0.0086 | 0.0003 | -28.5125 | <0.0001 | -0.0092 | -0.0080 |
| BMI per unit below 18.5 | -0.0199 | 0.0031 | -6.4609 | <0.0001 | -0.0260 | -0.0139 |
| Years since 2009 | 0.0173 | 0.0002 | 95.2821 | <0.0001 | 0.0169 | 0.0176 |
| Age | 0.0026 | 0.0000 | 82.2403 | <0.0001 | 0.0025 | 0.0027 |
| season_sin | -0.1151 | 0.0007 | -155.7826 | <0.0001 | -0.1165 | -0.1136 |
| season_cos | -0.1052 | 0.0007 | -149.7457 | <0.0001 | -0.1066 | -0.1039 |
| season_sin² | -0.0147 | 0.0007 | -20.9502 | <0.0001 | -0.0161 | -0.0133 |
| season_cos² | 0.0009 | 0.0007 | 1.2735 | 0.2028 | -0.0005 | 0.0024 |

**Model Summary:**

• **Dependent variable:** log_vitamin_d

• **R-squared:** 0.254

• **Adjusted R-squared:** 0.254

• **F-statistic:** 11,301 (p < 0.0001)

This table reports the results of a multivariable OLS regression model predicting serum 25(OH)D levels in adult LHS members with available demographic, socioeconomic, and BMI data. Independent variables include sex, ethnicity, socioeconomic status (SES), detailed BMI categories, age, year of test, and seasonal sinusoidal terms to model calendar trends. Negative coefficients indicate lower vitamin D levels associated with the corresponding category. The model explains 25.4% of the variance in log-transformed vitamin D levels.

### Supplementary Table 3. Regression model for vitamin D supplementation

| **Variable** | **Coefficient** | **Std. Error** | **t-Statistic** | **P-value** | **95% CI Lower** | **95% CI Upper** |
| --- | --- | --- | --- | --- | --- | --- |
| **vitd_kiu_m0** | 0.0127 | 0.0003 | 43.9128 | <0.0001 | 0.0121 | 0.0132 |
| **vitd_kiu_m1** | 0.0217 | 0.0003 | 68.0262 | <0.0001 | 0.0211 | 0.0223 |
| **vitd_kiu_m2** | 0.0216 | 0.0003 | 68.7724 | <0.0001 | 0.0210 | 0.0222 |
| **vitd_kiu_m3** | 0.0195 | 0.0003 | 64.0880 | <0.0001 | 0.0189 | 0.0201 |
| **vitd_kiu_m4** | 0.0148 | 0.0003 | 46.8482 | <0.0001 | 0.0142 | 0.0154 |
| **vitd_kiu_q1** | 0.0121 | 0.0003 | 39.3332 | <0.0001 | 0.0115 | 0.0127 |
| **vitd_kiu_q2** | 0.0094 | 0.0002 | 52.0134 | <0.0001 | 0.0091 | 0.0098 |
| **vitd_kiu_q3** | 0.0057 | 0.0002 | 31.4716 | <0.0001 | 0.0054 | 0.0061 |
| **vitd_kiu_q4** | 0.0041 | 0.0002 | 22.4462 | <0.0001 | 0.0038 | 0.0045 |
| **vitd_kiu_q5** | 0.0038 | 0.0002 | 20.0053 | <0.0001 | 0.0035 | 0.0042 |
| **vitd_kiu_q6** | 0.0032 | 0.0002 | 16.0337 | <0.0001 | 0.0028 | 0.0036 |
| **vitd_kiu_q7** | 0.0021 | 0.0002 | 9.9826 | <0.0001 | 0.0017 | 0.0025 |
| **vitd_kiu_y2** | 0.0016 | 0.0001 | 18.3022 | <0.0001 | 0.0014 | 0.0018 |
| **vitd_kiu_y3** | 0.0006 | 0.0001 | 6.6200 | <0.0001 | 0.0005 | 0.0008 |
| **vitd_kiu_y4** | 0.0007 | 0.0001 | 7.3420 | <0.0001 | 0.0005 | 0.0009 |

This table presents the results of a robust linear regression model estimating the difference between observed and predicted serum vitamin D levels as a function of vitamin D supplementation intensity over time. The dependent variable is the difference between measured serum vitamin D levels and the model-based estimate. Independent variables represent cumulative supplementation (in thousands of IU) over various time windows: monthly (vitd_kiu_m0 to m4), quarterly (q1 to q7), and yearly intervals (y2 to y4). All coefficients are statistically significant (p < 0.0001).

### Supplementary Table 4. List of predefined medical conditions used in the exploratory analysis phase in LHS to identify conditions associated with vitamin D deficiency

ATRIAL FIBRILLATION

ATRIAL FLUTTER

ADHD

ALCOHOLISM

ALZHEIMER

ANKYLOSING SPONDYLITIS

AORT ANEURYSM

ASTHMA

AUTISM

BEHCET

CA BREAST

CA COLON

CA KIDNEY

CA LUNG

CA METASTATIC

CA PANCREAS

CA PROSTATE

CA SKIN

CA SKIN BASAL

CA SKIN SQUAMOUS

CABG

CARDIOMYOPATHY

CELIAC

CEREBRAL BLEED

CHF

CIRRHOSIS

CKD

COAGULATION DISORDER

CONGENITAL HEART

CONNECT TISSUE DIS

COPD

CROHN'S DISEASE

CVA

DEAFNESS

DEMENTIA

DEPRESSION

DERMATOMYOSITIS

DIABETES INSIPIDUS

DIABETIC RETINOPATHY

DIALYSIS

DM

DM EYE DAMAGE

DM NEUROPATHY

DOWN

DRUG

DVT

DYSLIP

EATING DISORDER

EMBOLISM

ENDOCARDITIS

EPILEPSY

FIBROMYALGIA

FMF

G6PD DEF

GI BLEEDING

GI MALIGNANCY

GOUT

GRAVES

HASHIMOTO

HEMATOLOGIC MALIGNANCY

HEMIPLEGIA

HEPATITIS A

HEPATITIS AUTOIMMUNE

HEPATITIS B

HEPATITIS C

HEPATOCELLULAR CARCINOMA

HIDRADENITIS SUPPURATIVA

HIP FRACT

HTN

HYPERPARATHYROIDISM

HYPOTHYROIDISM

IBD

IDIOPATHIC PULMONARY FIBROSIS

IHD

IMMUNOSUPPRESSION

LEG AMPUTATION

LIVER DIS

LIVER DIS SEVERE

MACE

MELANOMA

MI

MULTIPLE SCLEROSIS

MYOCARDITIS

OSA

OSTEOART

OSTEOPOROSIS

PARKINSON

PCI

PULMONARY EMBOLISM

PERICARDITIS

PERNICIOUS ANEMIA

POLYMYOSITIS

PSORIASIS

PSORIAT ARTHRITIS

PTSD

PEPTIC ULCER DISEASE

PULM CONGESTION

PULM HTN

PULMONARY HYPERTENSION

PVD

RHEUMATOID ARTHRITIS

SCHIZOPHRENIA

SCLERODERMA

SIADH

SJOGREN

SLE

SOLID TUMOR

THROMBOSIS

TIA

ULCERATIVE COLITIS

ULCERATIVE PROCTITIS

URINARY MALIGNANCY

VALVE DISEASE

VALVE REPAIR

VALVE REPLACEMENT

ABDOMINAL PAIN

ACNE

ALLERGIC CONJUNCTIVITIS

ALLERGIC RHINITIS

ALLERGIC URTICARIA

ALLERGY FOOD/UNSPECIFIED

ALOPECIA AREATA

ANXIETY

ATOPIC DERMATITIS

BACK PAIN

BRONCHIOLITIS

BRONCHITIS

CANDIDA

CELLULITIS

CHEST PAIN

CLUSTER HEADACHE

COGNITIVE DISORDER

CONSTIPATION

CONTACT DERMATITIS

COUGH

COVID

DERMATITIS HERPETIFORMIS

DERMATOPHYTOSIS

DIARRHEA

ECZEMA DYSHIDROTIC

ENTEROBIASIS

ERYTHEMA MULTIFORME

ERYTHEMA NODOSUM

FALL

FECAL INCONTINENCE

FINGER OR HAND INJURY

FLATULENCE

FRACTURE

GASTRITIS

GASTROENTERITIS

GERD

GINGIVITIS

H PYLORI

HEADACHE

HELMINTHIASIS

HEMOPTYSIS

HEMORRHOIDS

HERPES SIMPLEX

HERPES ZOSTER

HYPOGLYCEMIA

IDIOPATHIC URTICARIA

IMPETIGO

INCONTINENCE

INFLUENZA

INGUINAL HERNIA

INSOMNIA

ITP

KETOACIDOSIS

LICHEN PLANUS

LUPUS DISCOID

MOLLUSCUM CONTAGIOSUM

NASAL POLYP

NOCTURNAL ENURESIS

PANCREATITIS

PEDICULOSIS

PEMPHIGOID

PEMPHIGUS

PERSONALITY DISORDERS

PITYRIASIS ROSEA

PNEUMONIA BACTERIAL

PNEUMONIA UNSPECIFIED

POLYMYALGIA RHEUMATICA

RASH

ROSACEA

SARCOIDOSIS

SCABIES

SEBORRHEIC DERMATITIS

SINUSITIS

SOMATISATION

STAPH INFECTION

TINEA VERSICOLOR

TOE OR LEG INJURY

TONSILLITIS

TREMOR

URTI

UTI

VERTIGO/DIZINESS

VIRAL INFECTION

VIRAL WART

VITILIGO

WHEEZING/STRIDOR

AMBLYOPIA

AMD

ASTIGMATISM

BLEPHARITIS

BLINDNESS

CATARACT

CONJUNCTIVITIS

CONJUNCTIVITIS CHRONIC

CORNEAL DISORDERS

CORNEAL EROSION

DISORDERS OPTIC NERVE AND VISUAL PATHWAYS

DISORDERS VITREOUS BODY

DRY EYE

ENTROPION OR TRICHIASIS

EPIPHORA

EYELID INFLAMMATION

EYELID OTHER

GLAUCOMA

HORDEOLUM OR CHALZION

HYPERMETROPIA

KERATITIS

KERATOCONUS

MYOPIA

ORBIT DISORDERS

PERIORBITAL CELLULITIS

PTERYGIUM

PTOSIS

REFRACTION DISORDERS

RETINAL DETACHMENT

### Supplementary Figure 1. Flowchart of Matched Vitamin D Cohort Construction and Matching

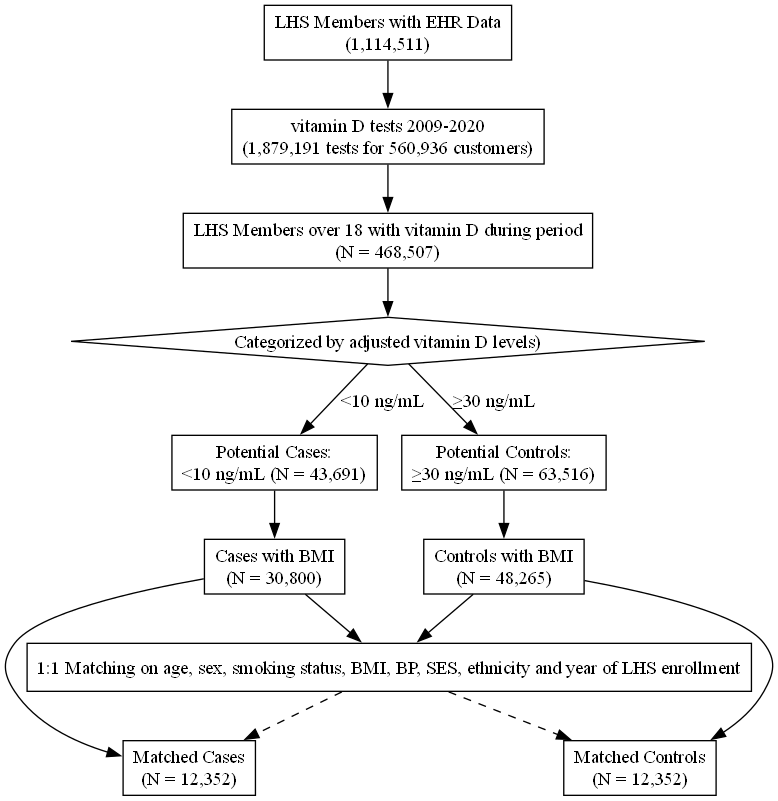

**Legend:**
Flowchart illustrating the construction of a matched cohort comparing individuals with severe vitamin D deficiency (<10 ng/mL) to those with sufficient levels (≥30 ng/mL) in the Leumit Health Services (LHS) database. Adults aged ≥18 years with at least one vitamin D test 2009-2020 were eligible. Individuals were categorized based on their calendar-adjusted serum 25(OH)D levels.
Only those with available BMI data were included for matching. A 1:1 nearest-neighbor matching was performed based on age, sex, smoking status, BMI, and BP categories, socioeconomic status (SES), ethnicity, and year of LHS enrollment yielding 12,352 matched pairs.

### Supplementary Figure 2. Sensitivity analyses performed with a 12-month lag

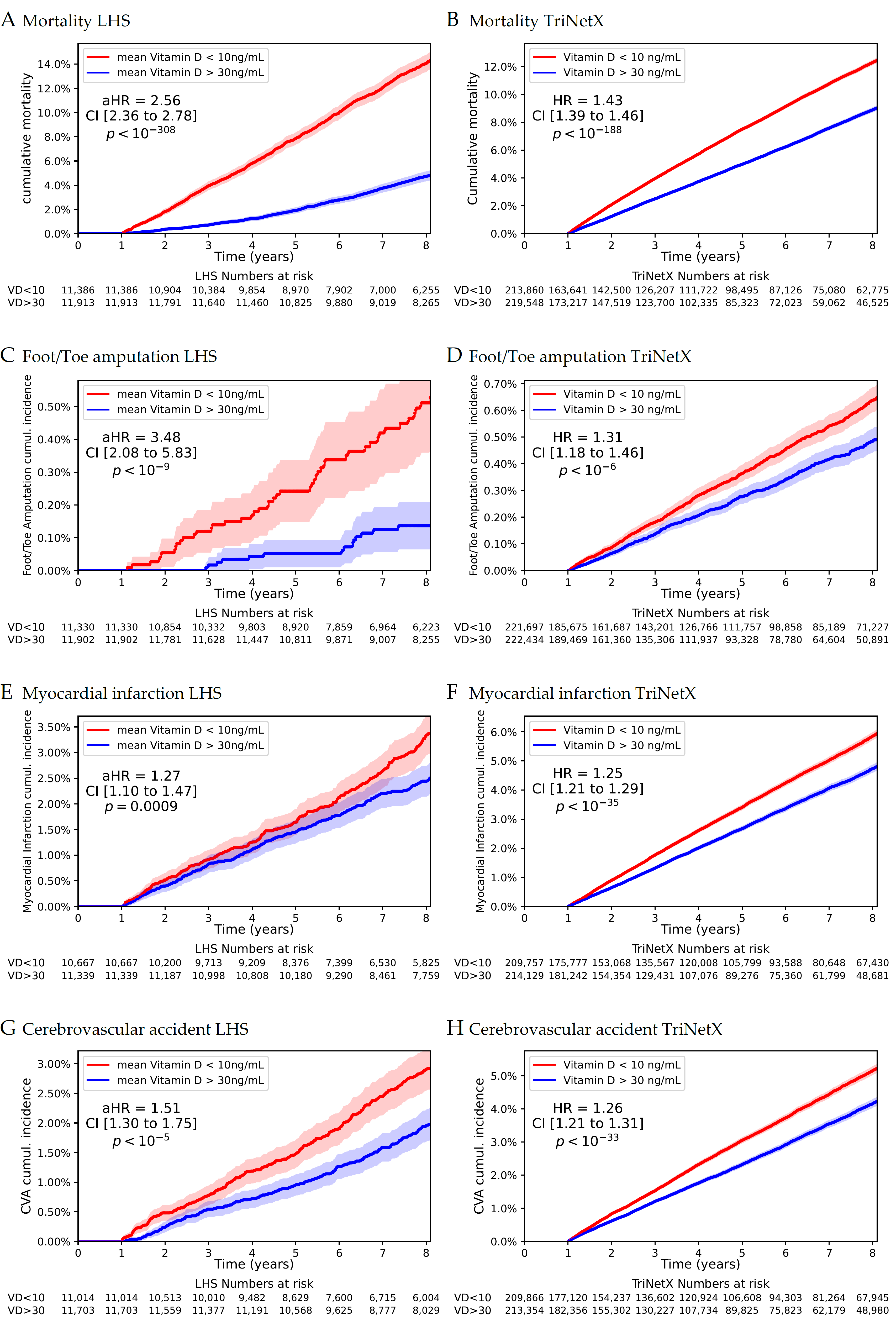

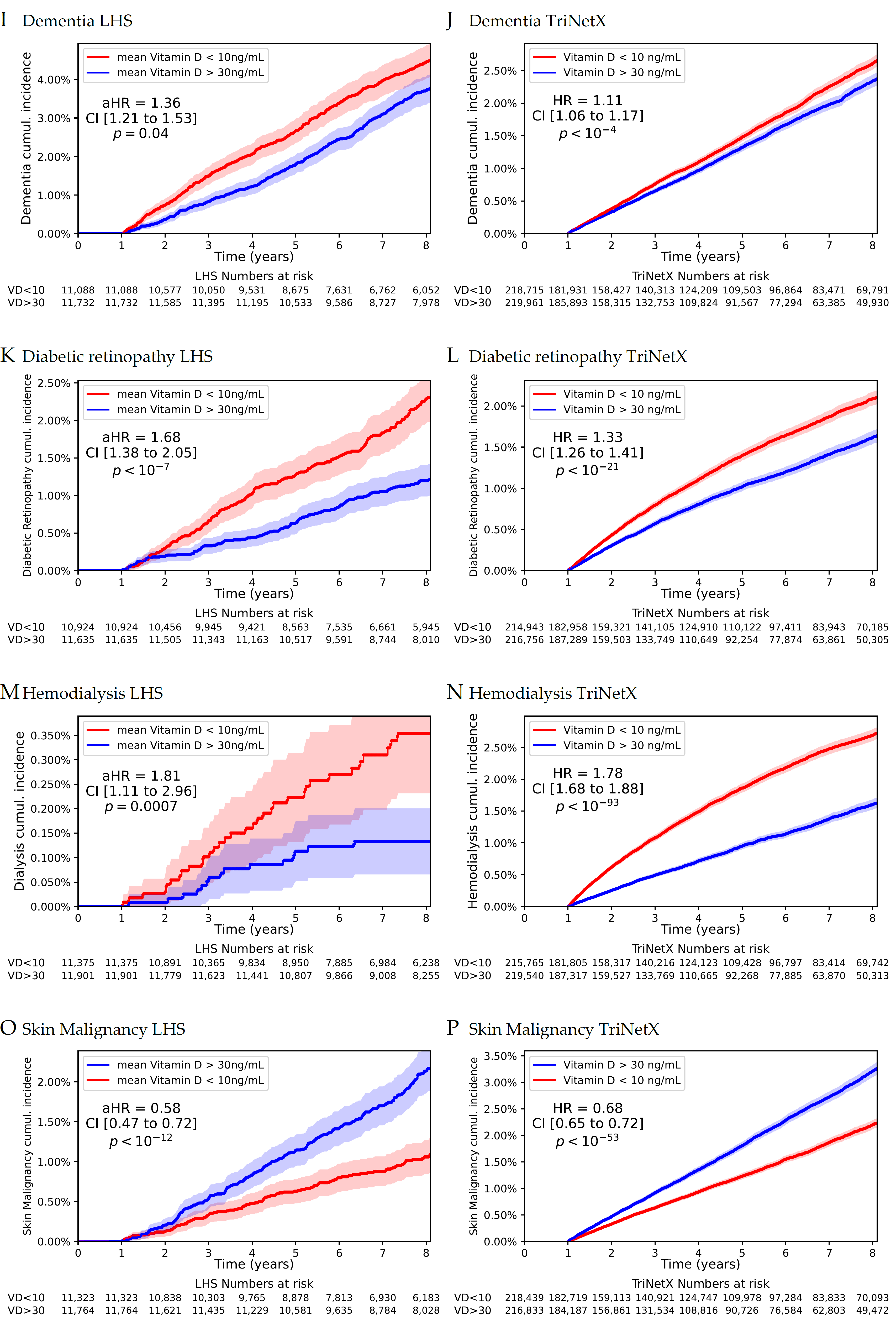
